# Dissecting the Genetic Relationship Between Severe Mental Disorders and Autoimmune Diseases

**DOI:** 10.1101/2025.06.22.25330080

**Authors:** Erik D Wiström, Julian Fuhrer, Alexey Shadrin, Vera Fominykh, Kevin S O’Connell, Piotr Jaholkowski, Nils Eiel Steen, Sara E Stinson, Pravesh Parekh, Oleksandr Frei, Linn Rødevand, Nadine Parker, Jan Haavik, Srdjan Djurovic, Anders M Dale, Ole A Andreassen, Olav B Smeland

## Abstract

Severe mental disorders have been linked to immune system dysfunction. While a genetic association between mental disorders and autoimmune diseases has been suggested, their genetic relationship remains incompletely understood. Utilizing a complementary set of statistical analyses, we conducted a comprehensive investigation of the genetic architecture between severe mental disorders (major depression (MD), bipolar disorder (BD), and schizophrenia (SCZ)) and seven autoimmune diseases (autoimmune thyroiditis, celiac disease, inflammatory bowel disease (IBD), multiple sclerosis, psoriasis, rheumatoid arthritis, and type 1 diabetes), involving a total of 667,518 cases from 10 genome-wide association studies. While MD was positively genetically correlated with five autoimmune diseases, BD and SCZ were only positively correlated with IBD, suggesting differences in the genetic signal shared with autoimmunity across these mental disorders. A considerable fraction of genetic variants influencing autoimmune diseases (range 17.1-88.4 %) were estimated to overlap with mental disorders; however, this constitutes only a minor part of genetic variants influencing the more polygenic mental disorders. Finally, we identified 172 genetic loci jointly affecting mental disorders and autoimmune diseases, implicating both lipid metabolism and TNF signaling. In conclusion, MD, BD, and SCZ have a small but distinct genetic overlap with autoimmune diseases, which may inform new possible immune targets for treatment in mental illness.

## Introduction

There is a growing interest in the relationship between severe mental disorders and the immune system, with evidence suggesting that immune processes may contribute to psychiatric pathophysiology (1). Major depression (MD), bipolar disorder (BD), and schizophrenia (SCZ) have all been linked to elevated markers of inflammation in serum (2,3), as well as in the cerebrospinal fluid (4). Further, there is a proposed link between infection and these mental disorders (5–7), mirroring the well-established link between infections and the onset of autoimmune disease (8).

A key area of current research is the potential involvement of autoimmune mechanisms in psychiatric conditions. Psychiatric symptoms such as depression, anxiety, and paranoia, are frequently observed in autoimmune diseases, including multiple sclerosis (MS), autoimmune thyroiditis (AITD), and inflammatory bowel disease (IBD) (9–11). Moreover, individuals with autoimmune diseases are at increased risk of developing MD, BD, and SCZ compared to the general population (12–14). Notably, this relationship appears to be bidirectional, whereby mental disorders similarly predispose individuals to the later onset of autoimmune disease (12–15).

Given that both groups of conditions are heritable, an important unresolved question is whether shared genetic mechanisms contribute to these observed associations (16,17). Several studies have reported positive genetic correlations between MD, BD, and SCZ and a variety of autoimmune diseases, such as IBD, psoriasis (PS), celiac disease (CeD), and MS (18–20). One study found that polygenic risk scores for SCZ predicted disease status in Crohn’s disease, type 1 diabetes (T1D), and rheumatoid arthritis (RA) (21). However, not all studies support a genetic association. A recent genome-wide association study (GWAS) on BD did not find any significant genetic correlations with autoimmune diseases (22), while another study found a small, but significant negative genetic correlation between SCZ and RA (23). Furthermore, while GWASs on MD, BD, and SCZ have identified genomic loci that are associated with both mental disorders and immune mechanisms (24–26), the specific genetic mechanisms that might underlie immunological abnormalities in severe mental disorders remain largely unknown. Clarifying these mechanisms might inform underlying psychopathology and lead to novel strategies for treatment and precision medicine development in psychiatry.

Advances in statistical analysis of large-scale GWAS data have enabled the study of genetic pleiotropy beyond genetic correlation, which has largely remained the standard approach for assessing genetic overlap between complex phenotypes using tools like linkage disequilibrium score regression (LDSC) (27). By comparison, the bivariate causal mixture model (MiXeR) and conjunctional false discovery rate (conjFDR) methods can quantify and identify the overlapping common genetic variants across traits regardless of the genetic correlation (28–30). These models are thus able to capture genetic pleiotropy even when single nucleotide polymorphisms (SNPs) exert mixed effect directions on the traits being studied and have been employed to explore pleiotropy across numerous complex phenotypes (31,32).

In this genetic study, we aimed to comprehensively dissect the genetic relationship between the severe mental disorders SCZ, BD, and MD and common autoimmune diseases, to pinpoint potentially shared molecular genetic mechanisms and to investigate the overlapping and distinct genetic architectures across these disorders. We analyzed the largest collection of GWAS summary statistics on these phenotypes to date, and applied the statistical methods LDSC (27), MiXeR (29), conjFDR (30), Multivariate Analysis of Genomic Annotation (MAGMA) (33), and Local Analysis of [co]Variant Association (LAVA) to investigate complementary aspects of genetic pleiotropy (34). For an overview of study design, see Figure 1.

**Fig. 1:**
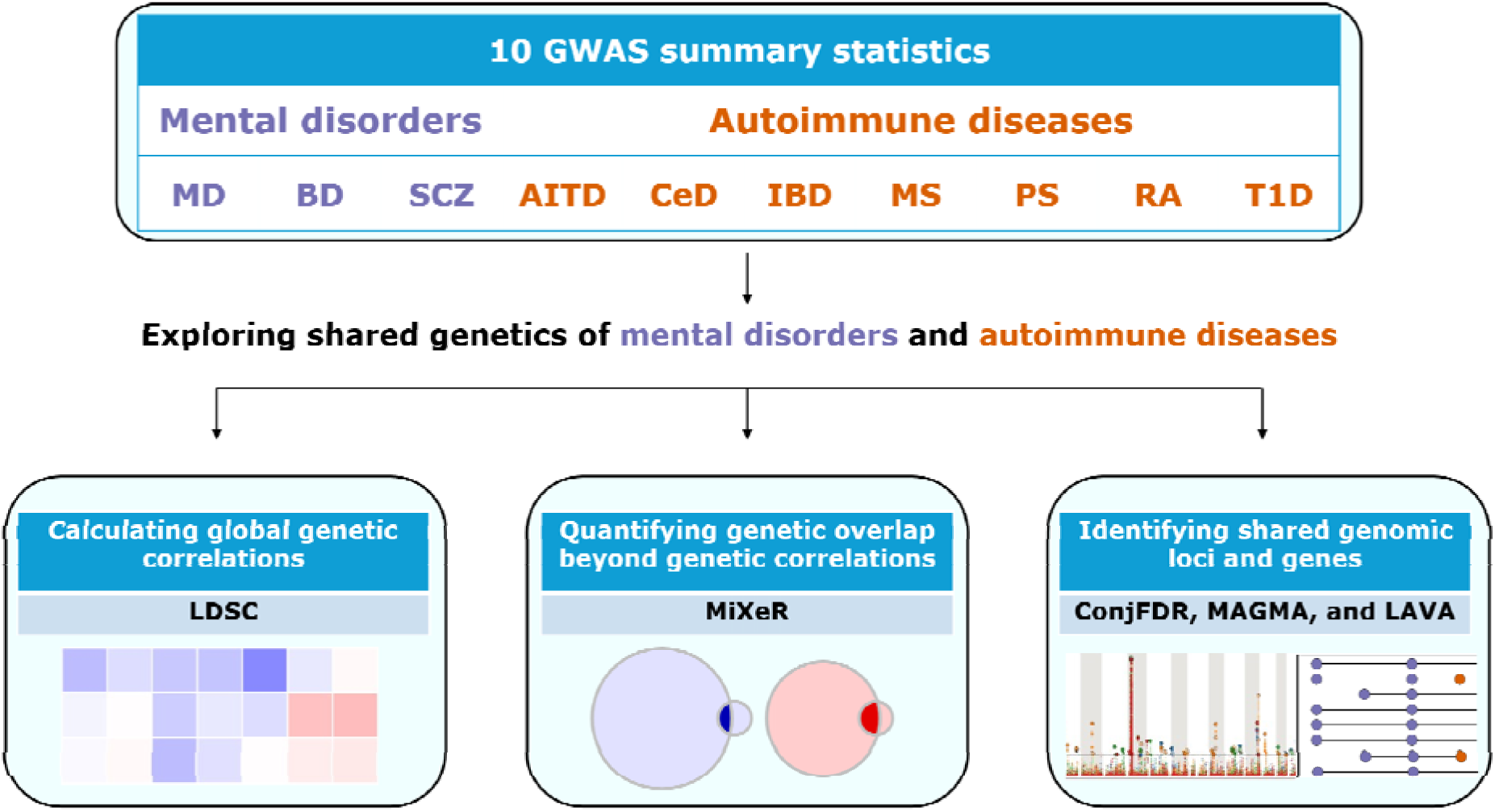
Study design overview. Overview of the genome-wide association study (GWAS) summary statistics and analyses performed in the study. Abbreviations of mental disorders: major depression (MD), bipolar disorder (BD), and schizophrenia (SCZ). Abbreviations of autoimmune diseases: autoimmune thyroiditis (AITD), celia disease (CeD), inflammatory bowel disease (IBD), multiple sclerosis (MS), psoriasis (PS), rheumatoid arthritis (RA), and type 1 diabetes (T1D). Abbreviations statistical methods: linkage disequilibrium score regression (LDSC), bivariate causal mixture model (MiXeR), conjunctional false discovery rate (conjFDR), Multivariate Analysis of Genomic Annotation (MAGMA), and Local Analysis of [co]Variant Association (LAVA).

## Material and Methods

### Participant samples and phenotypes

We collected publicly available GWAS summary statistics on the severe mental disorders MD, BD, and SCZ, totaling 524,978 cases (24–26). For autoimmune diseases, we focused on a broad set of conditions that affect different organ systems, with different involvement of the autoimmune-autoinflammatory axis (35,36), including AITD, CeD, IBD, MS, PS, RA, and T1D. We collected GWAS summary statistics from repositories and published studies. For CeD and PS, we performed GWAS meta-analyses utilizing data from UK Biobank, FinnGen, the Norwegian Mother, Father and Child Cohort Study (MoBa), and the Hordaland Health Study (HUSK) (37–40), to increase power for subsequent analyses, see Supplementary Note. Cases for autoimmune diseases collectively comprised 142,540 individuals (41–45). For an overview of cases and controls for each phenotype, see Table 1.

**Table 1.** Overview GWASs used in the study.

| Phenotype | Abbr. | N cases | N controls | GWAS source, PMID |
| --- | --- | --- | --- | --- |
| Major depression | MD | 412,305 | 1,588,397 | 37464041 (26) |
| Bipolar disorder | BD | 59,287 | 781,022 | 39843750 (24) |
| Schizophrenia | SCZ | 53,386 | 77,258 | 35396580 (25) |
| Autoimmune thyroiditis | AITD | 30,234 | 725,172 | 32581359 (41) |
| Celiac disease | CeD | 10,573 | 1,077,290 | UK Biobank, FinnGen (40),<br>MoBa (37), HUSK (38) |
| Inflammatory bowel disease | IBD | 25,042 | 34,915 | 28067908 (42) |
| Multiple sclerosis | MS | 14,802 | 26,703 | 31604244 (43) |
| Psoriasis | PS | 20,597 | 1,077,700 | UK Biobank, FinnGen (40),<br>MoBa (37), HUSK (38) |
| Rheumatoid arthritis | RA | 22,350 | 74,823 | 36333501 (44) |
| Type 1 diabetes | T1D | 18,942 | 501,638 | 34012112 (45) |

To avoid sample overlap in our conjFDR analyses, we removed cohorts from GWAS summary statistics on the mental disorder that were also included in the autoimmune GWASs. For more details on phenotype definition and participant samples, see Supplementary Note. Informed consent was gathered from each participant in the original studies. All participants had European ancestry, which ensured comparable LD patterns for follow-up analyses.

### Statistical analysis

#### Calculating genome-wide genetic correlations

We employed bivariate LDSC based on HapMap 3 SNPs to estimate genetic correlations between severe mental disorders and autoimmune diseases (27). To account for multiple testing, we applied the false discovery rate (FDR) procedure with a significance threshold of 0.05. The major histocompatibility complex (MHC) region was included, following the default procedure in these analyses. Given that MD exhibited positive genetic correlations with a greater number of autoimmune diseases than BD and SCZ, we conducted a sensitivity analysis assessing the potential impact of comorbid MD on these results using UK Biobank data. Specifically, we repeated the LDSC analyses between MD and the autoimmune diseases AITD, CeD, IBD, MS, and PS before and after excluding MD cases from the autoimmune disease UK biobank datasets, corrected for multiple comparisons using FDR.

#### Quantifying polygenic overlap beyond genetic correlations

We applied the Gaussian mixture models incorporated in MiXeR version 1.3 (29,46,47). First, we conducted a univariate MiXeR analysis calculating the polygenicity (number of trait-influencing variants) and the discoverability (average magnitude of effect sizes of trait-influencing variants) for each mental disorder and autoimmune disease. The model estimates the number of variants that collectively account for 90 % of the SNP heritability (*h^2^_SNP_*), excluding those with negligibly small effect sizes to ensure the robustness of the estimate. We then performed bivariate MiXeR to estimate the amount of overlap between trait-influencing variants affecting the phenotypes. We conducted 20 iterations of MiXeR analysis with 2 million random SNPs followed by random pruning at an r^2^ threshold of 0.8. The extended MHC region (genome build 19, location 26,000,000–34,000,000) was excluded due to its complex LD structure, following the default procedure.

We assessed model fit for univariate MiXeR by comparing Akaike information criterion (AIC) values between the MiXeR estimates and a reference model where all variants are assumed to be causal (infinitesimal model). A positive difference in AIC between MiXeR estimates and reference model indicates that the best-fitting MiXeR estimates are more accurate than the reference model. For bivariate MiXeR, the model fit was similarly evaluated by comparing the best-fitting MiXeR model against two reference models representing minimum and maximum possible overlap respectively. We present conditional quantile-quantile (Q-Q) plots and log-likelihood plots to visualize the stability of the fitness procedure.

#### Investigating local genetic correlations

We applied LAVA v. 1.3.8 (34) to identify local genetic correlations between mental disorders and autoimmune diseases across the 2,495 genomic regions of approximately 1 Mb in size, with the LD reference panel constructed based on individuals of European ancestry from the1000 Genomes phase 3 data. LAVA accounts for potential sample overlap using LDSC. After estimating the local *h_SNP_^2^* for each phenotype, we conducted pairwise local genetic correlation analysis for all regions with local *h_SNP_^2^* significantly different from zero. We utilized FDR correction to account for multiple testing. Furthermore, we compared the number of significant local genetic correlations within and outside of the extended MHC, defined according to the method’s recommendations (genome build 19, location 26,000,000–34,000,000).

#### Mapping shared genomic loci and genes

To increase statistical power to discover genetic loci that jointly affect severe mental disorders and autoimmune diseases, we used the conjFDR method (28,30). To control for spurious enrichment, we calculated average empirical distribution functions by randomly selecting one SNP in each LD block (r^2^1>10.1) over 500 iterations. To avoid bias in the FDR estimates, the extended MHC (genome build 19, location 25,119,106–33,854,733) and the chromosomal region 8p23.1 (location 7,200,000–12,500,000) were excluded before fitting the conditional cumulative distribution function, as this is the standard for the analysis (48).

We utilized the FUMA protocol to define genomic loci (49). For linking lead SNPs to the most likely causal genes, we used the open web platform Open Targets Genetics (https://genetics.opentargets.org/) (50). We performed gene expression and gene-set analysis applying FUMA, and Genotype-Tissue Expression (GTEx) dataset v8 on the genes identified by Open Targets, excluding lead SNPs located in the extended MHC region. We performed three sets of analyses with genes associated with MD, BD, and SCZ respectively. For more details, see Supplementary Note.

#### Identifying genome-wide significant shared genes

Using MAGMA version 1.08 (reference dataset 1,000 Genomes European panel) (33), we first identified likely causal genes for each GWAS and subsequently evaluated overlap of identified genes across psychiatric and autoimmune phenotypes. MAGMA calculates an association *P* value for each gene based on all SNPs mapped to the gene, accounting for gene size, number of SNPs within each gene, and LD between markers. We excluded the extended MHC from the analysis (genome build 19, location 26,000,000–34,000,000) as this is the default recommendation for the method. We set the significance threshold to 0.05 after Bonferroni correction for multiple testing.

## Results

### Global genetic correlations

Our LDSC analyses revealed different patterns of global genetic correlations among MD, BD, and SCZ with autoimmune diseases (Figure 2, Supplementary Table 1). While MD was significantly positively correlated with five autoimmune diseases (AITD, CeD, IBD, MS, and PS; r_g_ range 0.06-0.22), BD and SCZ were only positively correlated with IBD (0.10 and 0.13, respectively). Additionally, BD had significant negative correlations with RA and T1D (-0.12 and -0.13, respectively); no other genetic correlations were statistically significant after FDR correction. The genetic correlations within the two diagnostic categories are presented in Supplementary Figure 1 and Supplementary Table 2.

**Fig. 2:**
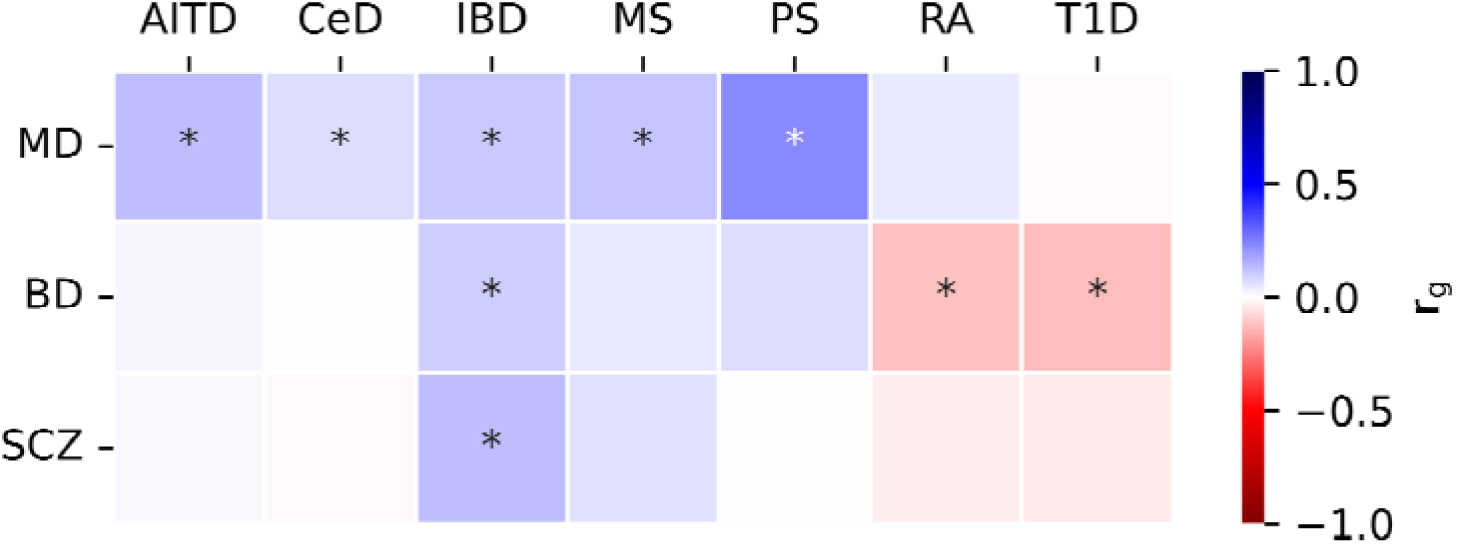
Global genetic correlations from LDSC analyses. Global genetic correlations observed between the psychiatric disorders major depression (MD), bipolar disorder (BD), and schizophrenia (SCZ), and the autoimmune diseases autoimmune thyroiditis (AITD), celiac disease (CeD), inflammatory bowel disease (IBD), multiple sclerosis (MS), psoriasis (PS), rheumatoid arthritis (RA), and type 1 diabetes (T1D). Colors represent r_g_ values. Statistically significant correlations, P <0.05 after FDR correction, are denoted by an asterisk.

To investigate whether the significant positive correlations between MD and autoimmune diseases could be driven by the presence of cases with comorbid MD among patients with autoimmune diseases, we repeated LDSC correlation analyses using UK Biobank data to generate autoimmune disease GWAS where we could exclude psychiatric diagnoses. Subsequently, we compared the LDSC results when including and excluding cases with comorbid MD using this dataset. While the LDSC analyses for AITD and MS became invalid due to non-significant *h_SNP_^2^* estimates, the other correlation estimates for CeD, IBD, and PS remained consistently positively correlated, though slightly attenuated (Pearson calculation= 0.9994, *P* value 0.02; Supplementary Table 3).

### Genetic overlap beyond genetic correlations

All univariate MiXeR analyses exhibited positive AIC values, indicating good model fit (Supplementary Table 4). We observed considerable differences in the estimated polygenicity across the two groups of conditions. The estimated number of trait-influencing variants ranged from 8.1K to 11.8K for severe mental disorders and from 0.13K to 0.73K for autoimmune diseases. The proportion of overlap of autoimmune disease variants was similar for most mental disorders. However, BD displayed a larger degree of overlap with MS, PS, RA, and T1D compared to MD and SCZ. Specifically, the proportion of shared trait-influencing variants for BD and autoimmune diseases ranged from 50.6 % of trait-influencing AITD variants to 88.4 % of trait-influencing T1D variants (Figure 3). Among bivariate analyses involving MD, T1D shared the lowest proportion of its trait-influencing variants with the disorder among the autoimmune diseases (34.6 %), while CeD shared the highest proportion of its variants (73.2 %). Bivariate analyses between SCZ and the autoimmune diseases showed overlap that ranged from 17.1 % of trait-influencing RA variants to 87.3 % of trait-influencing IBD variants. For bivariate MiXeR, comparisons of the best estimates to the minimum AIC yielded positive values for all analyses except for MD and PS, for which the analysis was unable to precisely estimate the number of shared variants (Supplementary Table 5, Supplementary Figure 2). For MiXeR estimates of required effective sample sizes to explain a specific percentage of *h_SNP_^2^* by genome-wide significant variants, see Supplementary Figure 3.

**Fig. 3:**
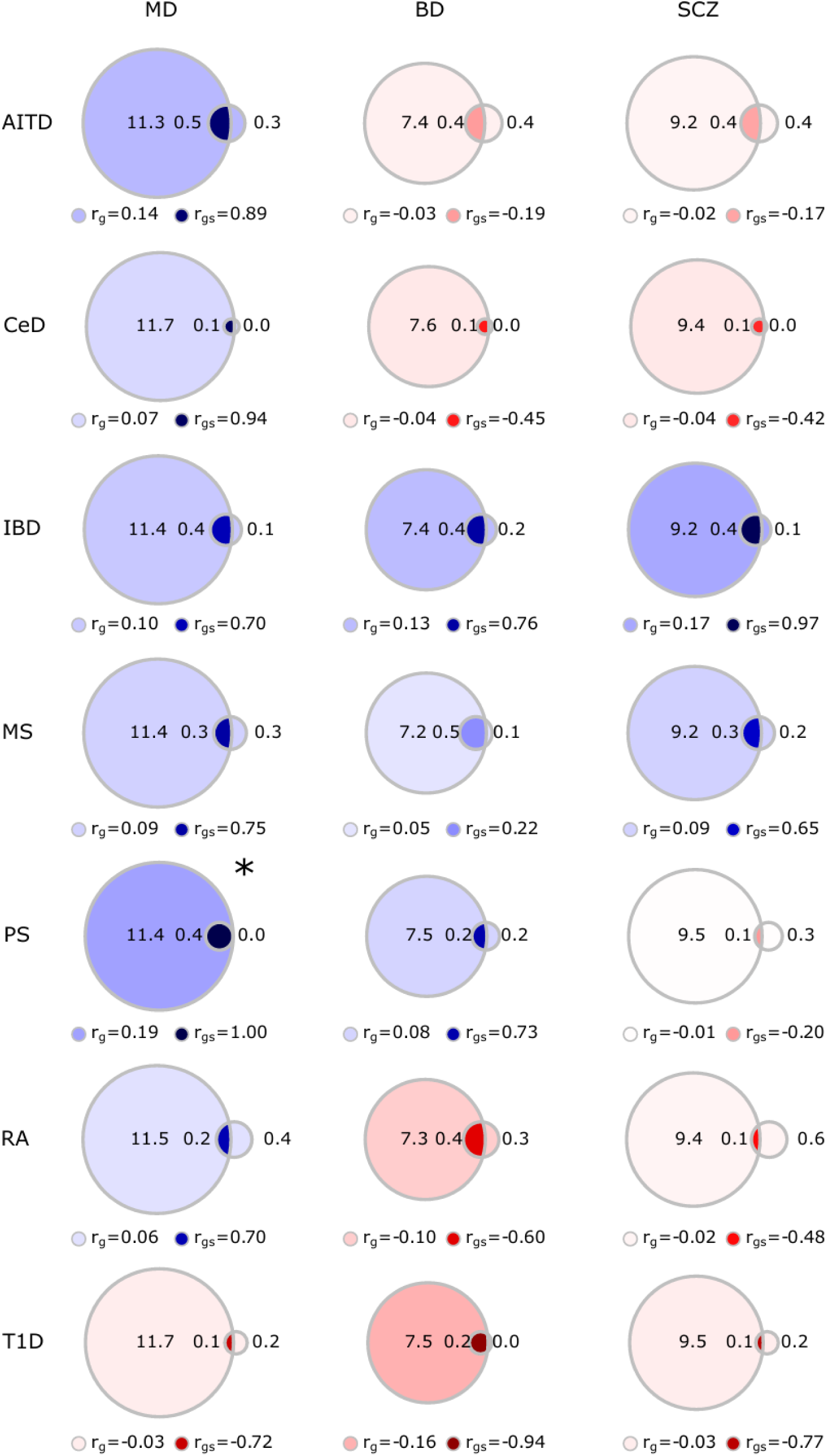
Genetic overlap estimated by MiXeR. Genetic overlap quantified by bivariate MiXeR analyses of the psychiatric disorders major depression (MD), bipolar disorder (BD), and schizophrenia (SCZ), and the autoimmune diseases autoimmune thyroiditis (AITD), celiac disease (CeD), inflammatory bowel disease (IBD), multiple sclerosis (MS), psoriasis (PS), rheumatoid arthritis (RA), and type 1 diabetes (T1D). Each bivariate MiXeR analysis is illustrated with the psychiatric disorder as the circle on the left, and the autoimmune disease as the circle on the right. Numbers within and adjacent to the circles indicate an estimate of trait-influencing variants in the thousands. Below are estimates of genetic correlations between the phenotypes, as well as genetic correlations of the shared variants. *In the MD and PS analysis, comparisons of the best estimate to the minimum Akaike information criterion (AIC) yielded a negative value, meaning that the analysis was unable to precisely estimate the number of shared variants.

### Local genetic correlations

In line with the MiXeR findings, we identified multiple genomic regions exhibiting significant local genetic correlations between severe mental disorders and autoimmune diseases using LAVA, irrespective of the global genetic correlations (Supplementary Table 6). For all mental disorders, we identified more regions with local correlations outside the MHC (105 for MD, 93 for BD, 144 for SCZ) than within the MHC (11 for MD, 14 for BD, 19 for SCZ) (Supplementary Figure 4), despite the well-established role of this locus in both psychiatric and autoimmune conditions (16,24–26). We also identified the most pleiotropic genomic hotspots, with 18 regions displaying significant local correlations involving at least three conditions (Supplementary Table 7). Among these, 12 were located within the extended MHC.

### Shared genetic loci and genome-wide significant genes

#### Shared genetic loci

Conditional Q-Q plots demonstrated cross-trait enrichment between mental disorders and autoimmune diseases (Supplementary Figure 5). Utilizing conjFDR, we identified 47 unique genetic loci associated with MD and at least one autoimmune disease, 14 of which were not found in the original GWAS, 38 unique loci associated with BD, 22 of which were not found in the original GWAS, and 87 unique loci associated with SCZ, 40 of which were not found in the original GWAS (Figure 4 and Supplementary Tables 8-28). Each conjFDR analysis between a mental disorder and an autoimmune disease implicated the extended MHC region. Outside of the MHC, 27 loci overlapped with two mental disorders, while only three loci were associated with all mental disorders.

**Fig. 4:**
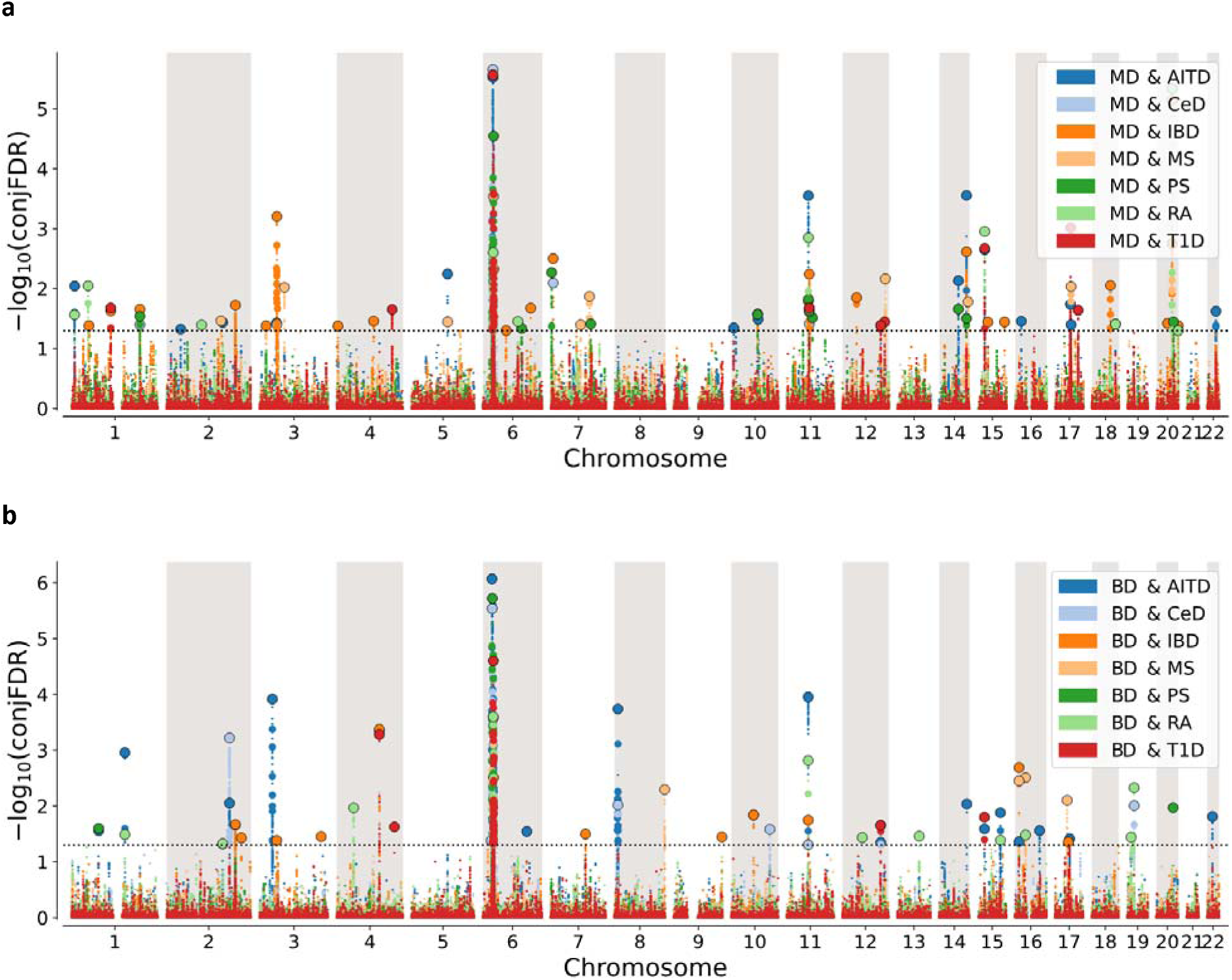
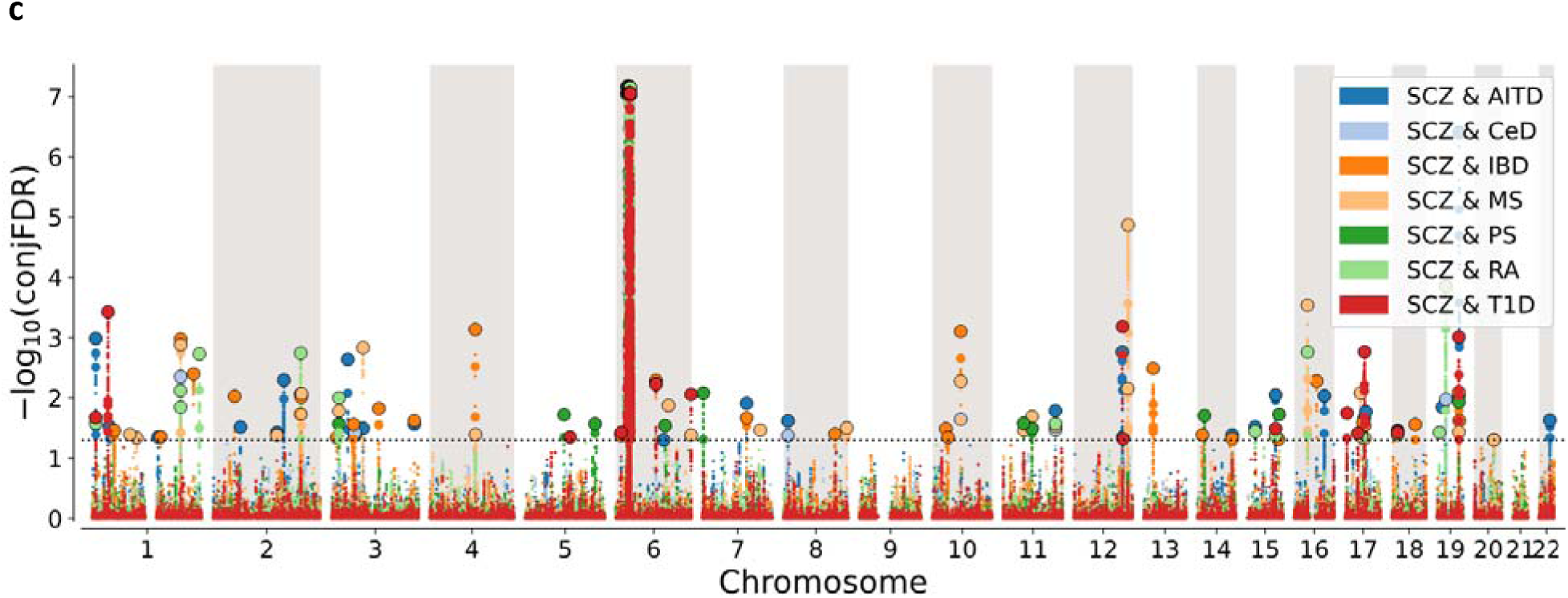
Miami plots of shared genome-wide significant loci identified by conjFDR. Conjunctional FDR (conjFDR) Manhattan plots illustrate genetic loci which major depression (MD) (a), bipolar disorder (BD) (b), and schizophrenia (SCZ) (c) share with the autoimmune diseases autoimmune thyroiditis (AITD), celiac disease (CeD), inflammatory bowel disease (IBD), multiple sclerosis (MS), psoriasis (PS), rheumatoid arthritis (RA), and type 1 diabetes (T1D). The x-axis represents chromosomal position, and the y-axis represents -log_10_ transformed FDR P values. Statistically significant loci at conjFDR <0.05 are encircled in black.

#### Mapped genes and enrichment analyses

For all identified loci, we identified the most likely causal gene using Open Targets. The MD-implicated genes were significantly differentially expressed (upregulated and downregulated) in the substantia nigra and frontal cortex regions of the brain, and they were downregulated in the liver and stomach. BD-implicated genes were not significantly differentially expressed in any specific tissue. SCZ-implicated genes were differentially expressed in the anterior cingulate cortex and the amygdala, as well as being downregulated in several other brain regions. For all tissue expression results, see Supplementary Figure 6. Gene-set enrichment analysis of MD-implicated genes highlighted significant involvement in the Gene Ontology (GO) biological processes “response to tumor necrosis factor (TNF)” and “central nervous system development” (Supplementary Table 9). For BD or SCZ associated genes, there was no significant enrichment in GO-related processes.

#### Shared genome-wide significant genes

We identified 78 genome-wide significant protein-coding genes shared between psychiatric and autoimmune conditions according to MAGMA (Supplementary Table 30). For an overview of the genes implicated by at least three different phenotypes, see Figure 5. The Forkhead Box P1 (*FOXP1*) gene was the only gene implicated by four disorders: MD, SCZ, IBD, and MS.

**Fig. 5:**
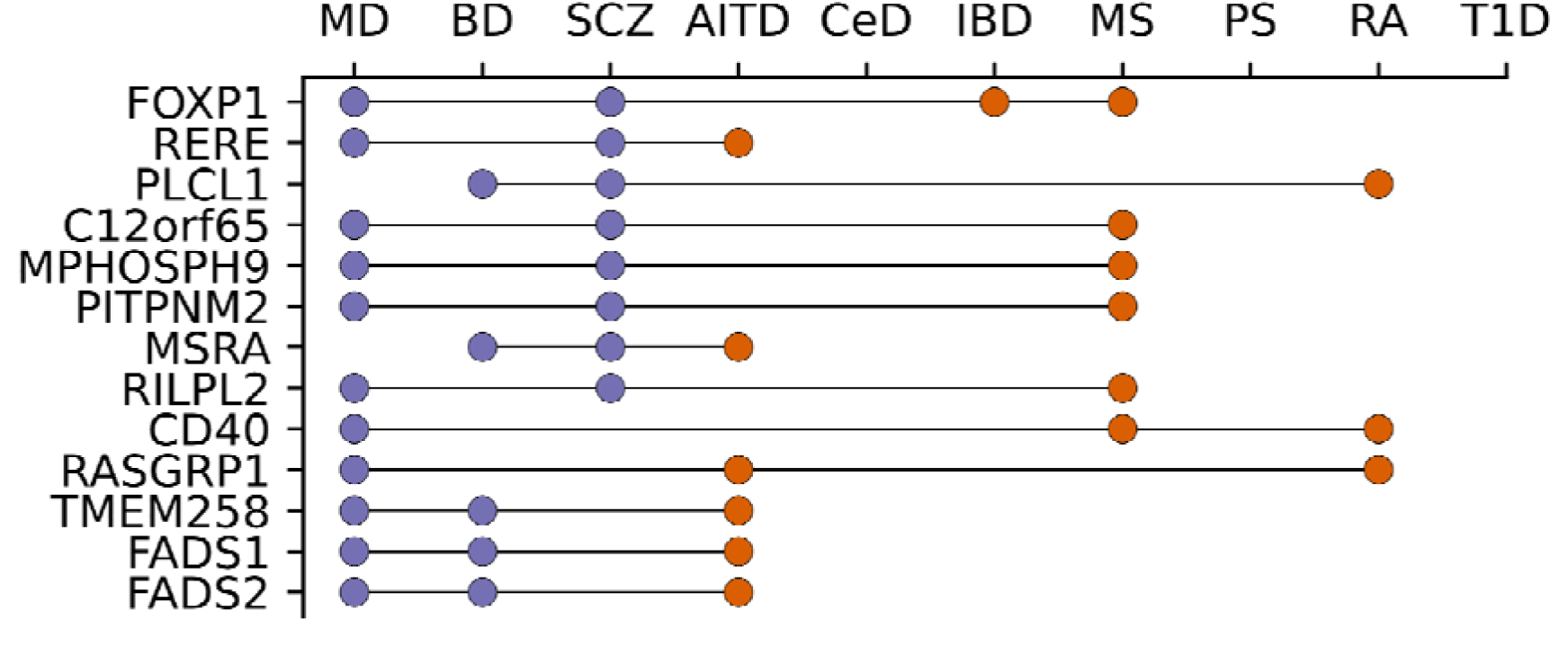
Shared genes according to MAGMA. Genes identified by MAGMA in three or more phenotypes after Bonferroni correction, including at least one of the psychiatric disorders major depression (MD), bipolar disorder (BD), and schizophrenia (SCZ), and at least one of the autoimmune diseases autoimmune thyroiditis (AITD), celiac disease (CeD), inflammatory bowel disease (IBD), multiple sclerosis (MS), psoriasis (PS), rheumatoid arthritis (RA), and type 1 diabetes (T1D).

## Discussion

In this genetic study, we employed a wide array of statistical methods to investigate the shared common variant architecture of the severe mental disorders MD, BD, and SCZ, and seven common autoimmune diseases. To our knowledge, this study represents the largest genomic investigation of this topic to date, involving a total of 667,518 cases from 10 GWAS datasets. Using LDSC, we found that MD has weak but significant positive genetic correlations with five autoimmune diseases: AITD, CeD, IBD, MS, and PS. In contrast, SCZ and BD were positively correlated with IBD only, indicating differences in the genetic signal shared with autoimmunity across these mental disorders. Furthermore, MiXeR revealed that a significant proportion of trait-influencing variants affecting autoimmune diseases also overlapped with mental disorders, even in the absence of significant genetic correlations between the phenotypes. Finally, we identified a large number of shared loci and genes, implicating both neurobiological and immunological mechanisms. Overall, the study provides new insights into the link between severe mental disorders and autoimmunity, suggesting potentially shared molecular genetic mechanisms that might inform new approaches to diagnosis and treatment of mental illness.

The LDSC analyses demonstrated weaker correlations between mental disorders and autoimmune diseases compared to the large correlations observed within the two diagnostic groups (Supplementary Figure 1), replicating well-established results in the literature (35,51). The finding that MD is more strongly correlated with autoimmune diseases diverges from previous LDSC studies based on smaller GWAS datasets, which have suggested comparable genetic correlations between the psychiatric and autoimmune conditions (18). Our sensitivity analyses indicated that comorbid MD among patients with autoimmune diseases does not explain this relationship, as the genetic correlations were only slightly attenuated after excluding individuals with a history of depression from the autoimmune datasets.

Among the autoimmune diseases, IBD distinguished itself as the only condition genetically correlated to all three mental disorders, with positive correlations from 0.10 to 0.13, in line with recent research from our group (19). A proposed link for a bidirectional relationship between severe mental disorders and IBD is the brain-gut axis, potentially mediated by the hypothalamus-pituitary-adrenal axis (52). In the most recent GWAS on BD, single-cell enrichment analysis across non-brain mouse tissues identified significant enrichment in the enteroendocrine cells of the large intestine and delta cells of the pancreas (24). Another noteworthy LDSC finding was the significant negative genetic correlations between BD and both RA and T1D, despite previous studies showing an increased prevalence of BD in both autoimmune diseases (53,54). This further emphasizes the complex genetic relationship between BD and autoimmune diseases, indicating that individuals with genetic risk of BD are at increased risk of IBD but lower risk of RA and T1D. Further research is needed to explain these negative associations.

The high estimated polygenicity of MD, BD, and SCZ is consistent with other complex brain and behavioral traits, such as general cognitive ability and neuroticism (55,56). Conversely, the lower polygenicity of autoimmune diseases is in the lower end of polygenicity estimates demonstrated for complex human disorders (19,57), akin to immunological biomarkers such as white blood cell counts and C-reactive protein level (31,32). Regardless of whether there were significant global genetic correlations between pairs of mental and autoimmune conditions, bivariate MiXeR estimated that more than half of the trait-influencing variants linked to autoimmune diseases were also linked to psychiatric disorders in most pairwise comparisons. This aligns with recent research from our own group, indicating that a large number of common variants have pleiotropic small effects across multiple diseases from different organ systems, but with a mixture of effect directions (19,56,58). Identifying these shared SNPs could reveal novel targets for improved diagnostics and personalized treatment strategies (59). For instance, gene expression analyses have shown promise in predicting non-response to antidepressants for MD patients (60). Moreover, clinical studies suggest that a subgroup of individuals with depression and immune dysfunction may benefit from medications targeting the immune system (61,62).

Using conjFDR, we identified the 172 most strongly associated shared loci between mental disorders and autoimmune diseases, approximately half of which were not identified in the original mental disorder GWASs. Interestingly, more than one-third of the genetic loci identified were shared by more than two conditions, suggesting pleiotropic effects across the mental and autoimmune conditions. Likewise, 13 genes identified using MAGMA and 48 correlated regions identified using LAVA were associated with at least three phenotypes. One notable locus on chromosome 11q12.2 (top lead SNPs rs174544 and rs174564) was identified in eight separate conjFDR analyses. This locus, associated with MD, BD, and five autoimmune diseases, implicates the fatty acid desaturase (*FADS*) genes, *FADS1* and *FADS2*, both previously linked to BD (24). This locus was also implicated by two separate LAVA analyses, and the *FADS1* and *FADS2* genes were also significantly associated with MD, BD, and AITD according to MAGMA. The FADS enzymes play crucial roles in fatty acid biosynthesis (63). Recent investigations of the human plasma metabolome have implicated abnormal fatty acid concentrations in both mental illness and inflammatory processes (64), and phospholipid-related alterations in cell membranes have been hypothesized in mental disorders (65).

Another locus also linked to lipid metabolism, located on chromosome 2q33.1 (top lead SNP rs976180), was significant in six conjFDR analyses. Previously known to be associated with MD and BD (24,26), this locus implicates the Phospholipase C Like 1 (*PLCL1*) gene. Underscoring its importance, *PLCL1* was also significant in three separate conditions in MAGMA (BD, SCZ and RA), and has previously been linked to both RA and the development of SCZ (66,67). The PLCL1 protein works as a binding partner for protein phosphatase 1 and 2A, which play important roles in adipose tissue lipolysis, as well as other cellular functions, including glycogen metabolism and regulation of interferon production (68–70). Another locus linked to protein phosphatase 1 was located on chromosome 11q13.1 (top lead SNP rs1783521), implicated in all seven conjFDR analyses for MD. This locus is associated with the Protein Phosphatase 1 Regulatory Inhibitor Subunit 14B (*PPP1R14B*) gene, which encodes an inhibitor of the serine/threonine phosphatase (71). Supporting its relevance, the locus also overlapped with a significant LAVA region.

Functional analysis of genes associated with MD and autoimmune diseases implicated the GO biological process “central nervous system development”, a known association for severe mental disorders and immune factors (1,59). Furthermore, the GO biological pathway “response to TNF” was also linked to the MD genes, despite the exclusion of the MHC region from this analysis, where the *TNF* gene itself is located. TNF, along with members of its superfamily, has important functions in the regulation of both innate and adaptive immune responses, and dysregulation of TNF signaling is linked to several autoimmune diseases (72). This finding is particularly notable given that the *TNF* gene is a drug target for infliximab, which has shown therapeutic potential in subgroups of patients with MD (62,73). Among the genes implicated in the TNF response pathway, two are targets for existing drugs according to the Open Targets Platform, though they have not been tested for treatment of MD; CD40 molecule (*CD40)* and TRAF3 Interacting Protein 2 (*TRAF3IP2*). *CD40* encodes a receptor expressed on B-cells which plays key roles in the adaptive immunity, among others by interacting with CD40 ligand on activated T-cells, thereby promoting B-cell differentiation and proliferation (74). *TRAF3IP2* is known to be associated with psoriasis (75), and encodes a protein which has regulatory effects on the homeostasis of B-cells by attenuating CD40 signaling (76). Underscoring its relevance, MAGMA identified *CD40* as genome-wide significant in three conditions. Also linked to TNF signaling was *FOXP1*, which was genome-wide significant in four conditions (MD, SCZ, IBD, and MS) according to MAGMA. Additionally, *FOXP1* was implicated by a genomic locus on chromosome 3p13 identified in three of our conjFDR analyses and one LAVA analysis. *FOXP1* encodes a transcription factor linked to stem cell differentiation in the bone marrow and early B cell development (77,78).

### Limitations

Our study has limitations that warrant consideration. Due to the intricate LD structure of the MHC, we did not have the ability to precisely map out shared genetic variants in this area with the statistical methods available. This is particularly notable given that the MHC has a critical role in autoimmunity and a strong link to mental disorders (1,16). Nevertheless, our LAVA analyses showed that, across all pairwise comparisons between mental disorders and autoimmune diseases, most shared genetic regions with local correlations resided outside of this locus. LD structures outside of the MHC may also have confounded our conjFDR results, as the lead SNPs assumed to be causal might merely be in close LD with the true causal variants. Similarly, SNP associations identified by conjFDR may not represent shared genetic signals, but distinct variants in close proximity to each other. Another limitation is our investigation’s restriction to participants of European ancestry to ensure LD compatibility with genetic references. This ancestry restriction possibly limits the generalizability of our findings to other populations. Potential bias in research participation might also be a concern, for instance SCZ encompasses a wide spectrum of severity and functional outcome that could skew study representation (79). Lastly, while we took measures to eliminate known sample overlap, we cannot exclude the possibility that some individuals participated in multiple studies, which could potentially confound the conjFDR results.

## Conclusions

This comprehensive genetic investigation shows that while severe mental disorders and autoimmune diseases have distinctly different genetic architectures, they also exhibit notable overlap, involving hundreds of common genetic variants with a mixture of effect directions. Our study highlights several genes that warrant further examination, particularly genes involved in lipid metabolism and TNF signaling.

## Acknowledgements

We would like to thank all the participants included in this study for making this research possible. We also thank the MVP, UK Biobank, IMSGC, PGC, Finngen, HUSK and MoBa for making their data available for us. All of the analyses were performed on Tjeneste for Sensitive Data (TSD), owned by the University of Oslo, operated and developed by the TSD service group, IT-Department (USIT). Analyses were also performed on resources provided by UNINETT Sigma2—the National Infrastructure for High Performance Computing and Data Storage, Norway (NS9666S). We want to acknowledge the participants and investigators of the FinnGen study. This research has been conducted using the UK Biobank Resource under Application Number 27412.

This study includes data from the MoBa conducted by the Norwegian Institute of Public Health. The MoBa is supported by the Norwegian Ministry of Health and Care Services and the Ministry of Education and Research. We are grateful to all the participating families in Norway who take part in this on-going cohort study. We thank the Norwegian Institute of Public Health (NIPH) for generating high-quality genomic data. This research is part of the HARVEST collaboration, supported by the Research Council of Norway (229624). We further thank the Center for Diabetes Research, the University of Bergen for providing genotype data and performing quality control and imputation of the data funded by the ERC AdG project SELECTionPREDISPOSED, Stiftelsen Kristian Gerhard Jebsen, Trond Mohn Foundation, the Research Council of Norway, the Novo Nordisk Foundation, the University of Bergen, and the Western Norway Health Authorities.

We are thankful for funding from the Research Council of Norway [grants 223273, 248778, 324252, 326813, 324499, 334920, 344121], the South-Eastern Norway Regional Health Authority [grant 2022–073, 2022-087], the European Union’s Horizon 2020 Research and Innovation Programme [grant 84776; CoMorMent, grant 964874; RealMent, Marie Skłodowska-Curie Actions grant 801133 (Scientia fellowship)], KG Jebsen Stiftelsen, the European Economic Area and Norway [grants #EEA-RO-NO-2018– 0573], and by the National Institutes of Health [grants 5R01MH124839–02 (PGC4), U24DA041123; R01AG076838; U24DA055330, OT2HL161847].

## Competing interests

Dr. Ole A. Andreassen has received speaker fees from Lundbeck, Janssen, Otsuka, Lilly, and Sunovion and is a consultant to Cortechs.ai. and Precision Health. Dr. Anders M. Dale is Founding Director, holds equity in CorTechs Labs, Inc. (DBA Cortechs.ai), and serves on its Board of Directors and Scientific Advisory Board. Dr. Dale is the President of J. Craig Venter Institute (JCVI) and is a member of the Board of Trustees of JCVI. He is an unpaid consultant for Oslo University Hospital. Dr. Oleksander Frei is a consultant for Precision Health. The other authors declare no competing interests.

## Data availability statement

GWAS summary statistics are publicly available at GWAS catalogue or available on request (International Multiple Sclerosis Genetic Consortium, HUSK, MoBa)

## Notes

### Author Declarations

All participants provided informed consent. The included studies had ethical approvals from institutional review boards or equivalent committees. The Norwegian Institutional Review Board for the South-East Norway Region has determined that no additional approval is required for the use of anonymized group-level data (ref. 2011/1980). Access to individual level UK Biobank data was obtained under the Application Number 27412. The establishment of MoBa and initial data collection was based on a license from the Norwegian Data Protection Agency and an approval from The Regional Committees for Medical and Health Research Ethics. The MoBa cohort is currently regulated by the Norwegian Health Registry Act. Participants in FinnGen provided informed consent for biobank research on basis of the Finnish Biobank Act. Alternatively, separate research cohorts, collected before the Finnish Biobank Act came into effect (in September 2013) and the start of FinnGen (August 2017) were collected on the basis of study-specific consent and later transferred to the Finnish biobanks after approval by Fimea, the National Supervisory Authority for Welfare and Health. The current HUSK study was approved by the Regional Committee for Medical and Health Research Ethics of Western Norway.

## References

1. Bennett FC, Molofsky AV. The immune system and psychiatric disease: a basic science perspective. Clinical and Experimental Immunology. 2019 Sep 1;197(3):294–307.

2. Ermakov EA, Melamud MM, Buneva VN, Ivanova SA. Immune System Abnormalities in Schizophrenia: An Integrative View and Translational Perspectives. Front Psychiatry. 2022 Apr 25;13.

3. Poletti S, Mazza MG, Benedetti F. Inflammatory mediators in major depression and bipolar disorder. Transl Psychiatry. 2024 Jun 8;14(1):1–13.

4. Orlovska-Waast S, Köhler-Forsberg O, Brix SW, Nordentoft M, Kondziella D, Krogh J, et al. Cerebrospinal fluid markers of inflammation and infections in schizophrenia and affective disorders: a systematic review and meta-analysis. Mol Psychiatry. 2019 Jun;24(6):869–87.

5. Yolken RH, Torrey EF. Are some cases of psychosis caused by microbial agents? A review of the evidence. Mol Psychiatry. 2008 May;13(5):470–9.

6. Wang X, Zhang L, Lei Y, Liu X, Zhou X, Liu Y, et al. Meta-Analysis of Infectious Agents and Depression. Sci Rep. 2014 Mar 31;4(1):4530.

7. Oliveira J, Oliveira-Maia AJ, Tamouza R, Brown AS, Leboyer M. Infectious and immunogenetic factors in bipolar disorder. Acta Psychiatrica Scandinavica. 2017;136(4):409–23.

8. Getts DR, Chastain EML, Terry RL, Miller SD. Virus infection, antiviral immunity, and autoimmunity. Immunological Reviews. 2013;255(1):197–209.

9. Sarısoy G, Terzi M, Gümüş K, Pazvantoğlu O. Psychiatric symptoms in patients with multiple sclerosis. General Hospital Psychiatry. 2013 Mar 1;35(2):134–40.

10. Leyhe T, Müssig K. Cognitive and affective dysfunctions in autoimmune thyroiditis. Brain, Behavior, and Immunity. 2014 Oct 1;41:261–6.

11. Kurina LM, Goldacre MJ, Yeates D, Gill LE. Depression and anxiety in people with inflammatory bowel disease. Journal of Epidemiology & Community Health. 2001 Oct 1;55(10):716–20.

12. Euesden J, Danese A, Lewis CM, Maughan B. A bidirectional relationship between depression and the autoimmune disorders – New perspectives from the National Child Development Study. PLOS ONE. 2017 Mar 6;12(3):e0173015.

13. Chen M, Jiang Q, Zhang L. The prevalence of bipolar disorder in autoimmune disease: a systematic review and meta-analysis. Annals of Palliative Medicine. 2021 Jan;10(1):35061– 361.

14. Jeppesen R, Benros ME. Autoimmune Diseases and Psychotic Disorders. Front Psychiatry. 2019 Mar 20;10:131.

15. Marrie RA, Walld R, Bolton JM, Sareen J, Walker JR, Patten SB, et al. Rising incidence of psychiatric disorders before diagnosis of immune-mediated inflammatory disease. Epidemiol Psychiatr Sci. 2019 Jun;28(3):333–42.

16. Pisetsky DS. Pathogenesis of autoimmune disease. Nat Rev Nephrol. 2023 Aug;19(8):509– 24.

17. Baselmans BML, Yengo L, van Rheenen W, Wray NR. Risk in Relatives, Heritability, SNP-Based Heritability, and Genetic Correlations in Psychiatric Disorders: A Review. Biological Psychiatry. 2021 Jan 1;89(1):11–9.

18. Tylee DS, Sun J, Hess JL, Tahir MA, Sharma E, Malik R, et al. Genetic correlations among psychiatric and immune-related phenotypes based on genome-wide association data. American Journal of Medical Genetics Part B: Neuropsychiatric Genetics. 2018;177(7):641– 57.

19. Smeland OB, Kutrolli G, Bahrami S, Fominykh V, Parker N, Hindley GFL, et al. The shared genetic risk architecture of neurological and psychiatric disorders: a genome-wide analysis. medRxiv. 2023 Sep 26;2023.07.21.23292993.

20. Levey DF, Stein MB, Wendt FR, Pathak GA, Zhou H, Aslan M, et al. Bi-ancestral depression GWAS in the Million Veteran Program and meta-analysis in> 1.2 million individuals highlight new therapeutic directions. Nature neuroscience. 2021;24(7):954–63.

21. Stringer S, Kahn RS, de Witte LD, Ophoff RA, Derks EM. Genetic liability for schizophrenia predicts risk of immune disorders. Schizophrenia Research. 2014 Nov 1;159(2):347–52.

22. Mullins N, Forstner AJ, O’Connell KS, Coombes B, Coleman JRI, Qiao Z, et al. Genome-wide association study of more than 40,000 bipolar disorder cases provides new insights into the underlying biology. Nat Genet. 2021 Jun;53(6):817–29.

23. Lee SH, Byrne EM, Hultman CM, Kähler A, Vinkhuyzen AA, Ripke S, et al. New data and an old puzzle: the negative association between schizophrenia and rheumatoid arthritis. International Journal of Epidemiology. 2015 Oct 1;44(5):1706–21.

24. O’Connell KS, Koromina M, van der Veen T, Boltz T, David FS, Yang JMK, et al. Genomics yields biological and phenotypic insights into bipolar disorder. Nature. 2025 Jan 22;1–12.

25. Trubetskoy V, Panagiotaropoulou G, Awasthi S, Braun A, Kraft J, Skarabis N, et al. Mapping genomic loci implicates genes and synaptic biology in schizophrenia. Nature. 2022 Apr;604(7906):502–8.

26. Adams MJ, Streit F, Meng X, Awasthi S, Adey BN, Choi KW, et al. Trans-ancestry genome-wide study of depression identifies 697 associations implicating cell types and pharmacotherapies. Cell. 2025 Feb 6;188(3):640–652.e9.

27. Bulik-Sullivan BK, Loh PR, Finucane HK, Ripke S, Yang J, Patterson N, et al. LD Score regression distinguishes confounding from polygenicity in genome-wide association studies. Nat Genet. 2015 Mar;47(3):291–5.

28. Andreassen OA, Thompson WK, Schork AJ, Ripke S, Mattingsdal M, Kelsoe JR, et al. Improved detection of common variants associated with schizophrenia and bipolar disorder using pleiotropy-informed conditional false discovery rate. PLoS Genet. 2013 Apr;9(4):e1003455.

29. Frei O, Holland D, Smeland OB, Shadrin AA, Fan CC, Maeland S, et al. Bivariate causal mixture model quantifies polygenic overlap between complex traits beyond genetic correlation. Nature communications. 2019;10(1):2417.

30. Smeland OB, Frei O, Shadrin A, O’Connell K, Fan CC, Bahrami S, et al. Discovery of shared genomic loci using the conditional false discovery rate approach. Hum Genet. 2020 Jan;139(1):85–94.

31. Hindley G, Kristian Drange O, Lin A, Kutrolli G, Shadrin AA, Parker N, et al. Cross-trait genome-wide association analysis of C-reactive protein level and psychiatric disorders. Psychoneuroendocrinology. 2023 Nov;157:106368.

32. Steen NE, Rahman Z, Szabo A, Hindley GFL, Parker N, Cheng W, et al. Shared Genetic Loci Between Schizophrenia and White Blood Cell Counts Suggest Genetically Determined Systemic Immune Abnormalities. Schizophrenia Bulletin. 2023 Sep 1;49(5):1345–54.

33. Leeuw CA de, Mooij JM, Heskes T, Posthuma D. MAGMA: Generalized Gene-Set Analysis of GWAS Data. PLoS Computational Biology. 2015 Apr 17;11(4):e1004219.

34. Werme J, van der Sluis S, Posthuma D, de Leeuw CA. An integrated framework for local genetic correlation analysis. Nat Genet. 2022 Mar;54(3):274–82.

35. Fominykh V, Shadrin AA, Jaholkowski P, Fuhrer J, Parker N, Wiström ED, et al. Mapping the genetic landscape of immune-mediated disorders: potential implications for classification and therapeutic strategies. Front Immunol. 16.

36. Breunig S, Lee YH, Karlson EW, Krishnan A, Lawrence JM, Schaffer LS, et al. Examining the Genetic Links between Clusters of Immune-mediated Diseases and Psychiatric Disorders. medRxiv. 2024 Jul 19;2024.07.18.24310651.

37. Magnus P, Birke C, Vejrup K, Haugan A, Alsaker E, Daltveit AK, et al. Cohort Profile Update: The Norwegian Mother and Child Cohort Study (MoBa). International Journal of Epidemiology. 2016 Apr 1;45(2):382–8.

38. Refsum H, Nurk E, David Smith A, Ueland PM, Gjesdal CG, Bjelland I, et al. The Hordaland Homocysteine Study: A Community-Based Study of Homocysteine, Its Determinants, and Associations with Disease1. The Journal of Nutrition. 2006 Jun 1;136(6):1731S–1740S.

39. Paltiel L, Anita H, Skjerden T, Harbak K, Bækken S, Kristin SN, et al. The biobank of the Norwegian Mother and Child Cohort Study – present status. Norsk Epidemiologi. 2014 Dec 22;24(1–2).

40. Kurki MI, Karjalainen J, Palta P, Sipilä TP, Kristiansson K, Donner KM, et al. FinnGen provides genetic insights from a well-phenotyped isolated population. Nature. 2023 Jan;613(7944):508–18.

41. Saevarsdottir S, Olafsdottir TA, Ivarsdottir EV, Halldorsson GH, Gunnarsdottir K, Sigurdsson A, et al. FLT3 stop mutation increases FLT3 ligand level and risk of autoimmune thyroid disease. Nature. 2020 Aug;584(7822):619–23.

42. de Lange KM, Moutsianas L, Lee JC, Lamb CA, Luo Y, Kennedy NA, et al. Genome-wide association study implicates immune activation of multiple integrin genes in inflammatory bowel disease. Nat Genet. 2017 Feb;49(2):256–61.

43. International Multiple Sclerosis Genetics Consortium. Multiple sclerosis genomic map implicates peripheral immune cells and microglia in susceptibility. Science. 2019 Sep 27;365(6460):eaav7188.

44. Ishigaki K, Sakaue S, Terao C, Luo Y, Sonehara K, Yamaguchi K, et al. Multi-ancestry genome-wide association analyses identify novel genetic mechanisms in rheumatoid arthritis. Nat Genet. 2022 Nov;54(11):1640–51.

45. Chiou J, Geusz RJ, Okino ML, Han JY, Miller M, Melton R, et al. Interpreting type 1 diabetes risk with genetics and single-cell epigenomics. Nature. 2021 Jun;594(7863):398–402.

46. Holland D, Frei O, Desikan R, Fan CC, Shadrin AA, Smeland OB, et al. Beyond SNP heritability: Polygenicity and discoverability of phenotypes estimated with a univariate Gaussian mixture model. PLOS Genetics. 2020 May 19;16(5):e1008612.

47. Meer D van der, Hindley G, Shadrin AA, Smeland OB, Parker N, Dale AM, et al. Mapping the Genetic Landscape of Psychiatric Disorders With the MiXeR Toolset. Biological Psychiatry. 2025 Feb 19;0(0).

48. Schwartzman A, Lin X. The effect of correlation in false discovery rate estimation. Biometrika. 2011 Mar 1;98(1):199–214.

49. Watanabe K, Taskesen E, van Bochoven A, Posthuma D. Functional mapping and annotation of genetic associations with FUMA. Nat Commun. 2017 Nov 28;8(1):1826.

50. Ghoussaini M, Mountjoy E, Carmona M, Peat G, Schmidt EM, Hercules A, et al. Open Targets Genetics: systematic identification of trait-associated genes using large-scale genetics and functional genomics. Nucleic acids research. 2021;49(D1):D1311–20.

51. Lee PH, Anttila V, Won H, Feng YCA, Rosenthal J, Zhu Z, et al. Genomic Relationships, Novel Loci, and Pleiotropic Mechanisms across Eight Psychiatric Disorders. Cell. 2019 Dec 12;179(7):1469–1482.e11.

52. Gracie DJ, Hamlin PJ, Ford AC. The influence of the brain–gut axis in inflammatory bowel disease and possible implications for treatment. The Lancet Gastroenterology & Hepatology. 2019 Aug 1;4(8):632–42.

53. Charoenngam N, Ponvilawan B, Ungprasert P. Patients with rheumatoid arthritis have a higher risk of bipolar disorder: A systematic review and meta-analysis. Psychiatry Research. 2019 Dec 1;282:112484.

54. Chen MH, Tsai SJ, Bai YM, Huang KL, Su TP, Chen TJ, et al. Type 1 diabetes mellitus and risks of major psychiatric disorders: A nationwide population-based cohort study. Diabetes & Metabolism. 2022 Jan 1;48(1):101319.

55. Karadag N, Hagen E, Shadrin AA, van der Meer D, O’Connell KS, Rahman Z, et al. Unraveling the shared genetics of common epilepsies and general cognitive ability. Seizure: European Journal of Epilepsy. 2024 Nov 1;122:105–12.

56. Hindley G, Frei O, Shadrin AA, Cheng W, O’Connell KS, Icick R, et al. Charting the Landscape of Genetic Overlap Between Mental Disorders and Related Traits Beyond Genetic Correlation. AJP. 2022 Nov;179(11):833–43.

57. Watanabe K, Stringer S, Frei O, Umićević Mirkov M, de Leeuw C, Polderman TJC, et al. A global overview of pleiotropy and genetic architecture in complex traits. Nat Genet. 2019 Sep;51(9):1339–48.

58. Smeland OB, Frei O, Dale AM, Andreassen OA. The polygenic architecture of schizophrenia — rethinking pathogenesis and nosology. Nat Rev Neurol. 2020 Jul;16(7):366–79.

59. Lee PH, Feng YCA, Smoller JW. Pleiotropy and Cross-Disorder Genetics Among Psychiatric Disorders. Biological Psychiatry. 2021 Jan 1;89(1):20–31.

60. Cattaneo A, Gennarelli M, Uher R, Breen G, Farmer A, Aitchison KJ, et al. Candidate Genes Expression Profile Associated with Antidepressants Response in the GENDEP Study: Differentiating between Baseline ‘Predictors’ and Longitudinal ‘Targets’. Neuropsychopharmacology. 2013 Feb;38(3):377–85.

61. Bialek K, Czarny P, Strycharz J, Sliwinski T. Major depressive disorders accompanying autoimmune diseases – Response to treatment. Progress in Neuro-Psychopharmacology and Biological Psychiatry. 2019 Dec 20;95:109678.

62. Arteaga-Henríquez G, Simon MS, Burger B, Weidinger E, Wijkhuijs A, Arolt V, et al. Low-grade inflammation as a predictor of antidepressant and anti-inflammatory therapy response in MDD patients: a systematic review of the literature in combination with an analysis of experimental data collected in the EU-MOODINFLAME consortium. Frontiers in psychiatry. 2019;10:458.

63. Merino DM, Ma DW, Mutch DM. Genetic variation in lipid desaturases and its impact on the development of human disease. Lipids Health Dis. 2010 Jun 18;9(1):63.

64. Julkunen H, Cichońska A, Tiainen M, Koskela H, Nybo K, Mäkelä V, et al. Atlas of plasma NMR biomarkers for health and disease in 118,461 individuals from the UK Biobank. Nat Commun. 2023 Feb 3;14(1):604.

65. Yao JK, van Kammen DP. Membrane phospholipids and cytokine interaction in schizophrenia. Int Rev Neurobiol. 2004;59:297–326.

66. Luo S, Li XF, Yang YL, Song B, Wu S, Niu XN, et al. PLCL1 regulates fibroblast-like synoviocytes inflammation via NLRP3 inflammasomes in rheumatoid arthritis. Advances in Rheumatology. 2022 Jul 11;62(1):25.

67. Notaras M, Lodhi A, Dündar F, Collier P, Sayles NM, Tilgner H, et al. Schizophrenia is defined by cell-specific neuropathology and multiple neurodevelopmental mechanisms in patient-derived cerebral organoids. Mol Psychiatry. 2022 Mar;27(3):1416–34.

68. Wies E, Wang MK, Maharaj NP, Chen K, Zhou S, Finberg RW, et al. Dephosphorylation of the RNA Sensors RIG-I and MDA5 by the Phosphatase PP1 Is Essential for Innate Immune Signaling. Immunity. 2013 Mar 21;38(3):437–49.

69. Okumura T, Harada K, Oue K, Zhang J, Asano S, Hayashiuchi M, et al. Phospholipase C-Related Catalytically Inactive Protein (PRIP) Regulates Lipolysis in Adipose Tissue by Modulating the Phosphorylation of Hormone-Sensitive Lipase. PLOS ONE. 2014 Jun 19;9(6):e100559.

70. Ceulemans H, Bollen M. Functional Diversity of Protein Phosphatase-1, a Cellular Economizer and Reset Button. Physiological Reviews. 2004 Jan;84(1):1–39.

71. Eto M, Karginov A, Brautigan DL. A Novel Phosphoprotein Inhibitor of Protein Type-1 Phosphatase Holoenzymes. Biochemistry. 1999 Dec 1;38(51):16952–7.

72. Croft M, Salek-Ardakani S, Ware CF. Targeting the TNF and TNFR superfamilies in autoimmune disease and cancer. Nat Rev Drug Discov. 2024 Dec;23(12):939–61.

73. Drevets WC, Wittenberg GM, Bullmore ET, Manji HK. Immune targets for therapeutic development in depression: towards precision medicine. Nat Rev Drug Discov. 2022 Mar;21(3):224–44.

74. Elgueta R, Benson MJ, De Vries VC, Wasiuk A, Guo Y, Noelle RJ. Molecular mechanism and function of CD40/CD40L engagement in the immune system. Immunological Reviews. 2009;229(1):152–72.

75. Ellinghaus E, Ellinghaus D, Stuart PE, Nair RP, Debrus S, Raelson JV, et al. Genome-wide association study identifies a psoriasis susceptibility locus at TRAF3IP2. Nat Genet. 2010 Nov;42(11):991–5.

76. Qian Y, Liu C, Hartupee J, Altuntas CZ, Gulen MF, Jane-wit D, et al. The adaptor Act1 is required for interleukin 17–dependent signaling associated with autoimmune and inflammatory disease. Nat Immunol. 2007 Mar;8(3):247–56.

77. Hu H, Wang B, Borde M, Nardone J, Maika S, Allred L, et al. Foxp1 is an essential transcriptional regulator of B cell development. Nat Immunol. 2006 Aug;7(8):819–26.

78. Li H, Liu P, Xu S, Li Y, Dekker JD, Li B, et al. FOXP1 controls mesenchymal stem cell commitment and senescence during skeletal aging. J Clin Invest. 127(4):1241–53.

79. Lang FU, Kösters M, Lang S, Becker T, Jäger M. Psychopathological long-term outcome of schizophrenia – a review. Acta Psychiatrica Scandinavica. 2013;127(3):173–82.

