## Supplementary Note for "Dissecting the Genetic Relationship Between Severe Mental Disorders and Autoimmune Diseases"

### Table of contents

Supplementary Figures……………………………………………………………………………………………………………….1

Supplementary Figure 1. Global genetic correlations from extended LDSC analyses……………………1

Supplementary Figure 2. Q-Q plots and log-likelihood plots from bivariate MiXeR analyses…………2

Supplementary Figure 3. GWAS Power plots estimated by MiXeR…………………………………………….9

Supplementary Figure 4. Loci identified by Local Analysis of [co]Variant Association (LAVA)………..10

Supplementary Figure 5. Q-Q plots from conjFDR analyses…………………………………………………….11

### Supplementary Figures

#### Supplementary Figure 1. Global genetic correlations from extended LDSC analyses

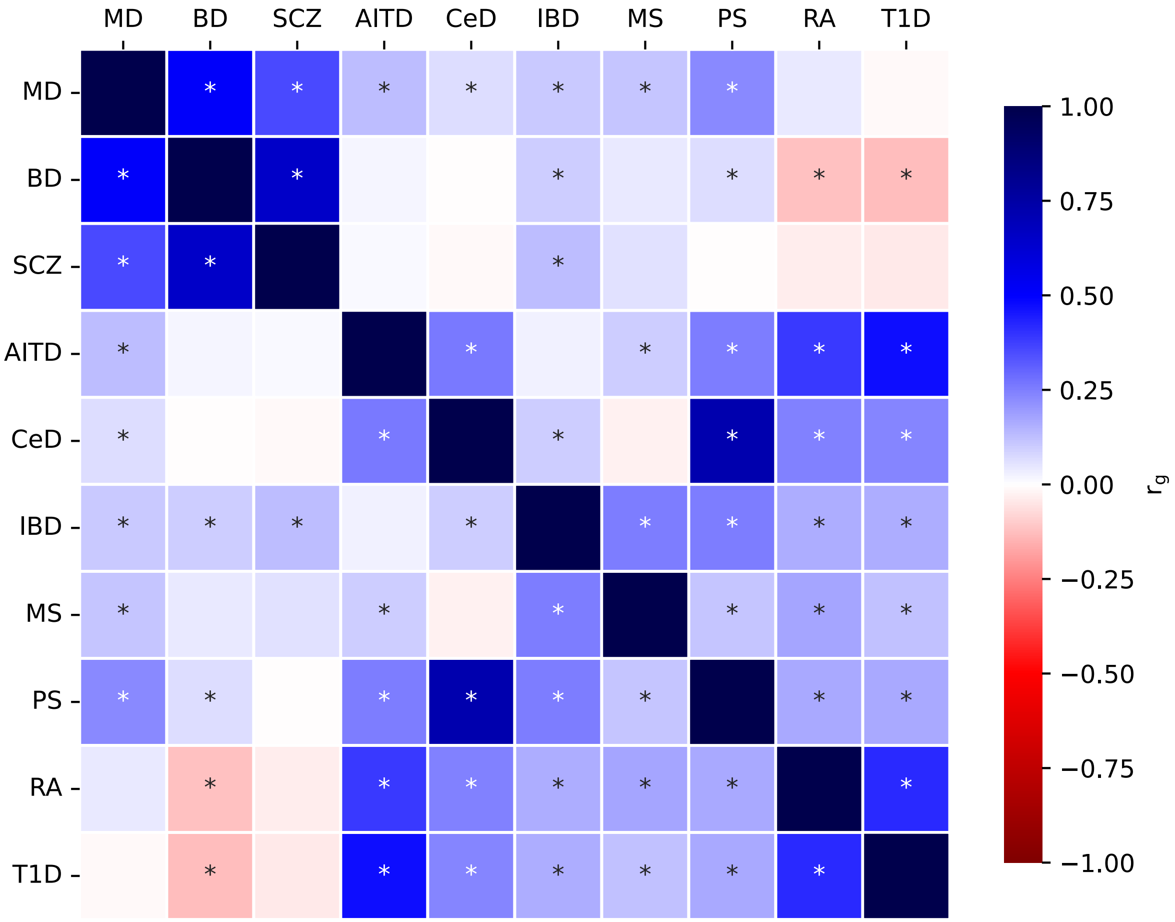

Global genetic correlations observed between major depression (MD), bipolar disorder (BD), schizophrenia (SCZ), autoimmune thyroiditis (AITD), celiac disease (CeD), inflammatory bowel disease (IBD), multiple sclerosis (MS), psoriasis (PS), rheumatoid arthritis (RA), and type 1 diabetes (T1D). Colors represent $r_g$ values. Statistically significant correlations, *P* <0.05 after FDR correction, are denoted by an asterisk.

**Supplementary Figure 2. Q-Q plots and log-likelihood plots from bivariate MiXeR analyses**

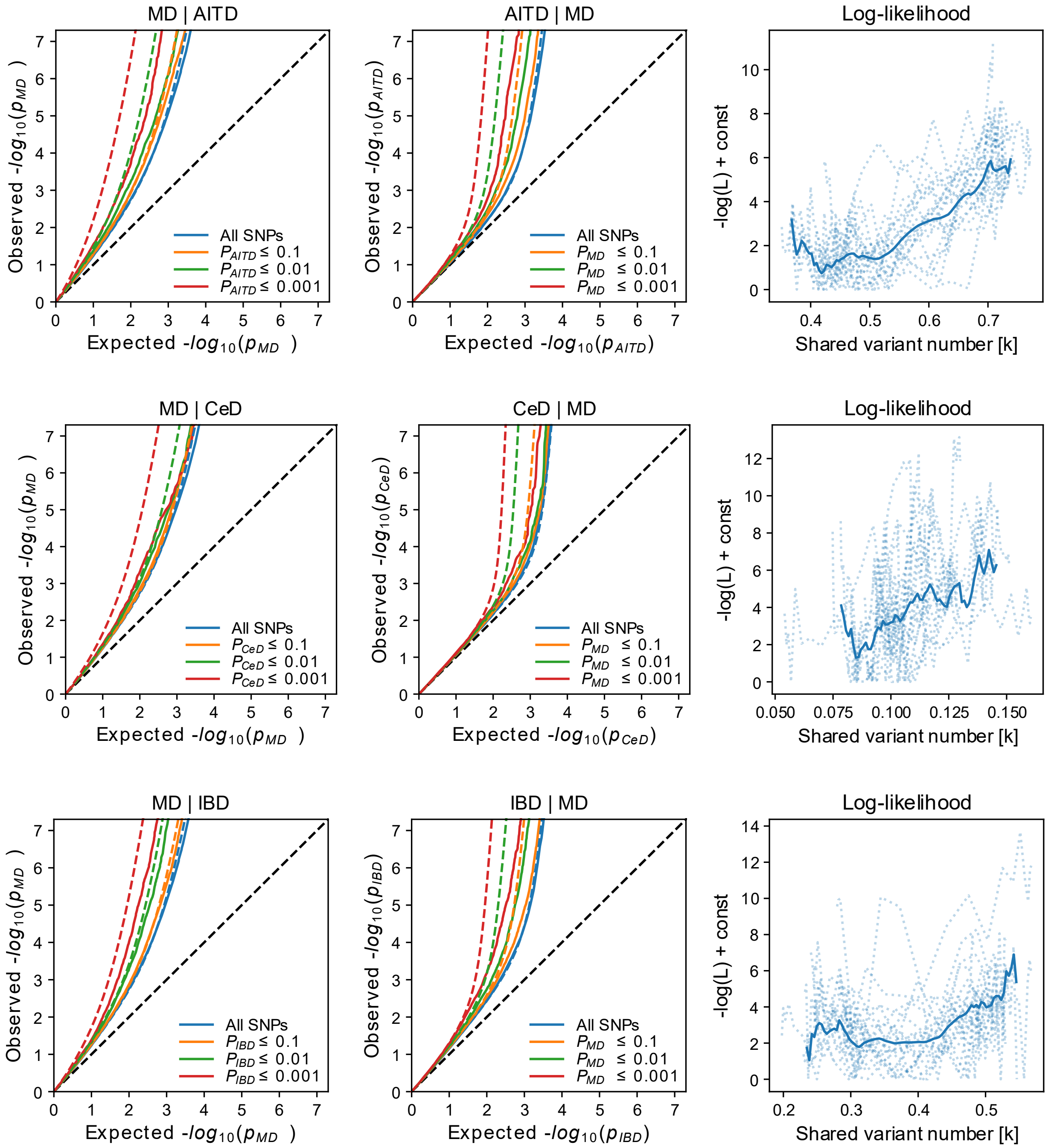

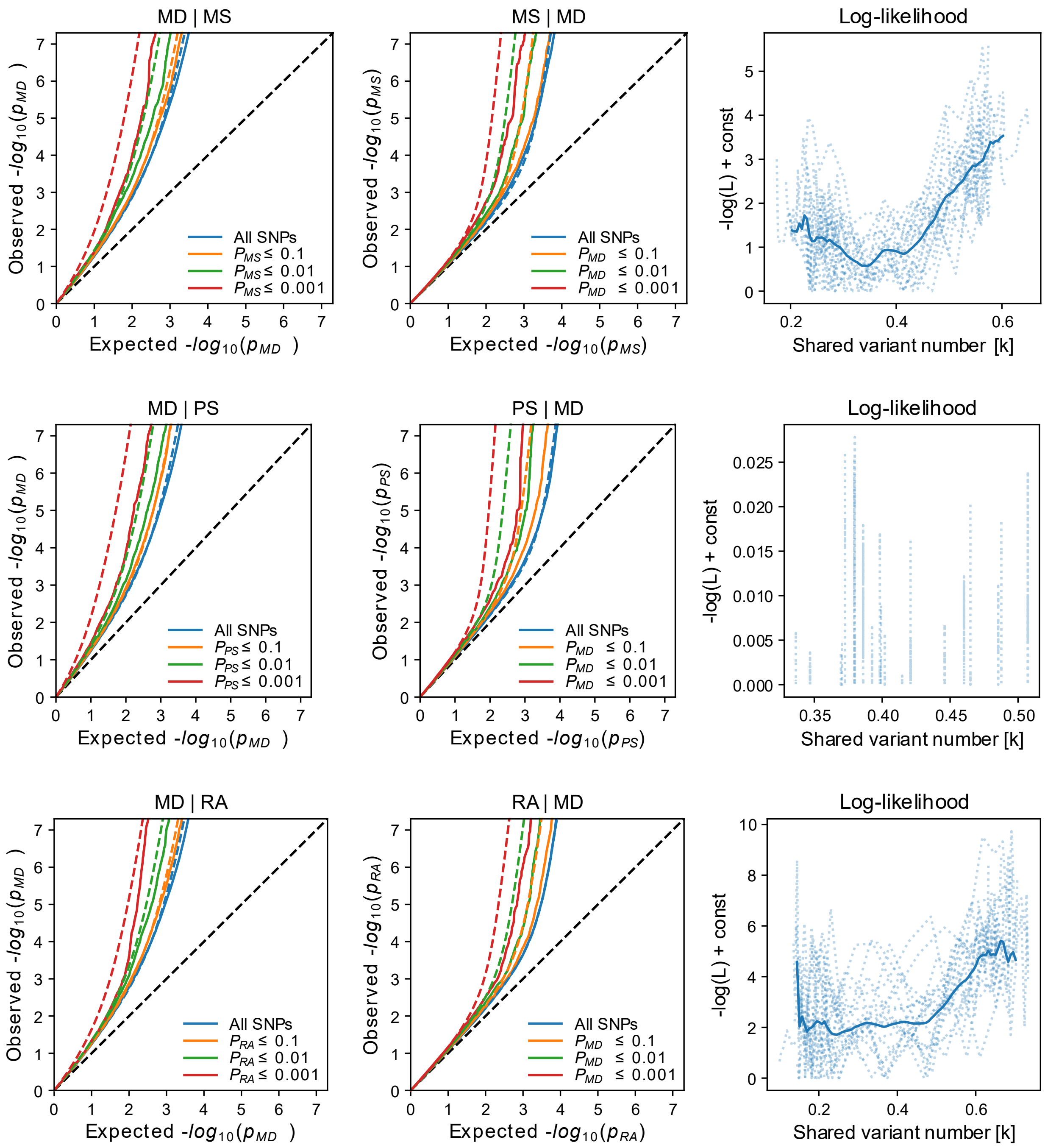

MD | MS
MS | MD
Log-likelihood
MD | PS
PS | MD
Log-likelihood
MD | RA
RA | MD
Log-likelihood
Observed -log10(pMD)
Observed -log10(pMS)
Observed -log10(pPS)
Observed -log10(pRA)
Expected -log10(pMD)
Expected -log10(pMS)
Expected -log10(pPS)
Expected -log10(pRA)
-log(L) + const
Shared variant number [k]
All SNPs
PMS ≤ 0.1
PMS ≤ 0.01
PMS ≤ 0.001
PMD ≤ 0.1
PMD ≤ 0.01
PMD ≤ 0.001
PPS ≤ 0.1
PPS ≤ 0.01
PPS ≤ 0.001
PRA ≤ 0.1
PRA ≤ 0.01
PRA ≤ 0.001

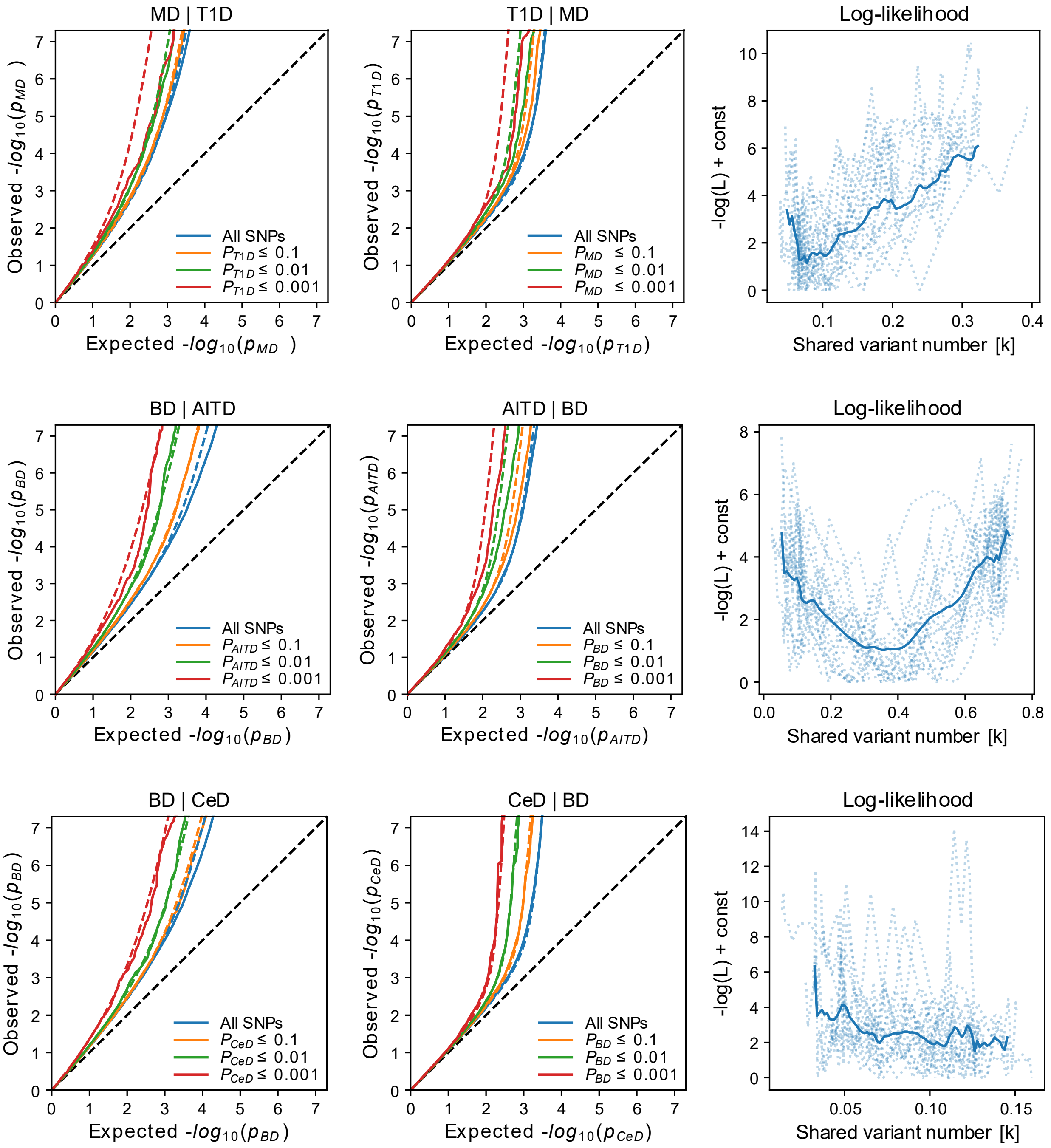
MD | T1D
T1D | MD
Log-likelihood
BD | AITD
AITD | BD
Log-likelihood
BD | CeD
CeD | BD
Log-likelihood
Observed -$log_{10}(p_{MD})$
Expected -$log_{10}(p_{MD})$
Observed -$log_{10}(p_{T1D})$
Expected -$log_{10}(p_{T1D})$
Observed -$log_{10}(p_{BD})$
Expected -$log_{10}(p_{BD})$
Observed -$log_{10}(p_{AITD})$
Expected -$log_{10}(p_{AITD})$
Observed -$log_{10}(p_{CeD})$
Expected -$log_{10}(p_{CeD})$
-log(L) + const
Shared variant number [k]
All SNPs
$P_{T1D} \leq 0.1$
$P_{T1D} \leq 0.01$
$P_{T1D} \leq 0.001$
$P_{MD} \leq 0.1$
$P_{MD} \leq 0.01$
$P_{MD} \leq 0.001$
$P_{AITD} \leq 0.1$
$P_{AITD} \leq 0.01$
$P_{AITD} \leq 0.001$
$P_{BD} \leq 0.1$
$P_{BD} \leq 0.01$
$P_{BD} \leq 0.001$
$P_{CeD} \leq 0.1$
$P_{CeD} \leq 0.01$
$P_{CeD} \leq 0.001$

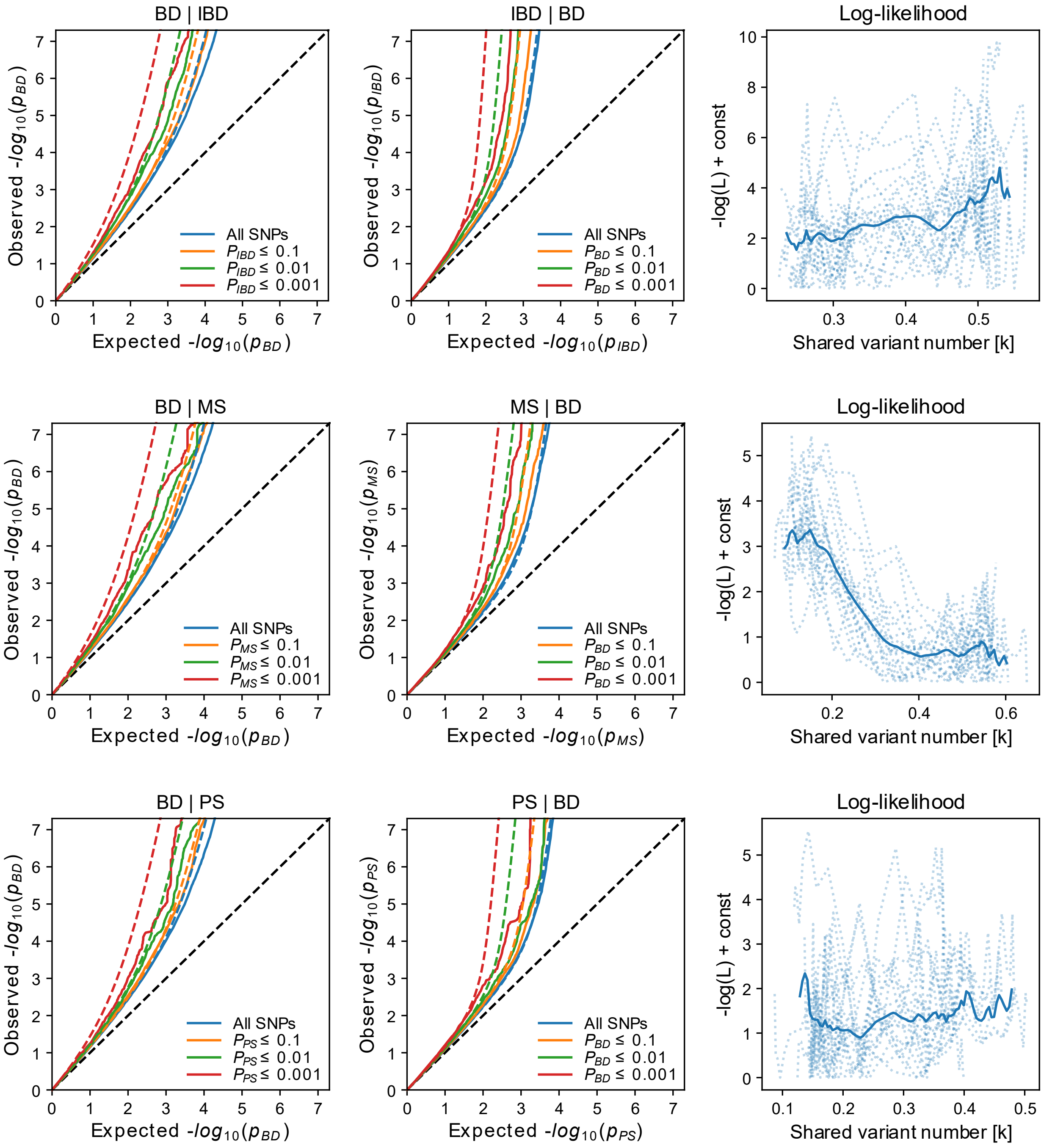

BD | IBD
IBD | BD
Log-likelihood
BD | MS
MS | BD
Log-likelihood
BD | PS
PS | BD
Log-likelihood
All SNPs
Observed -log10(p_BD)
Expected -log10(p_BD)
-log(L) + const
Shared variant number [k]

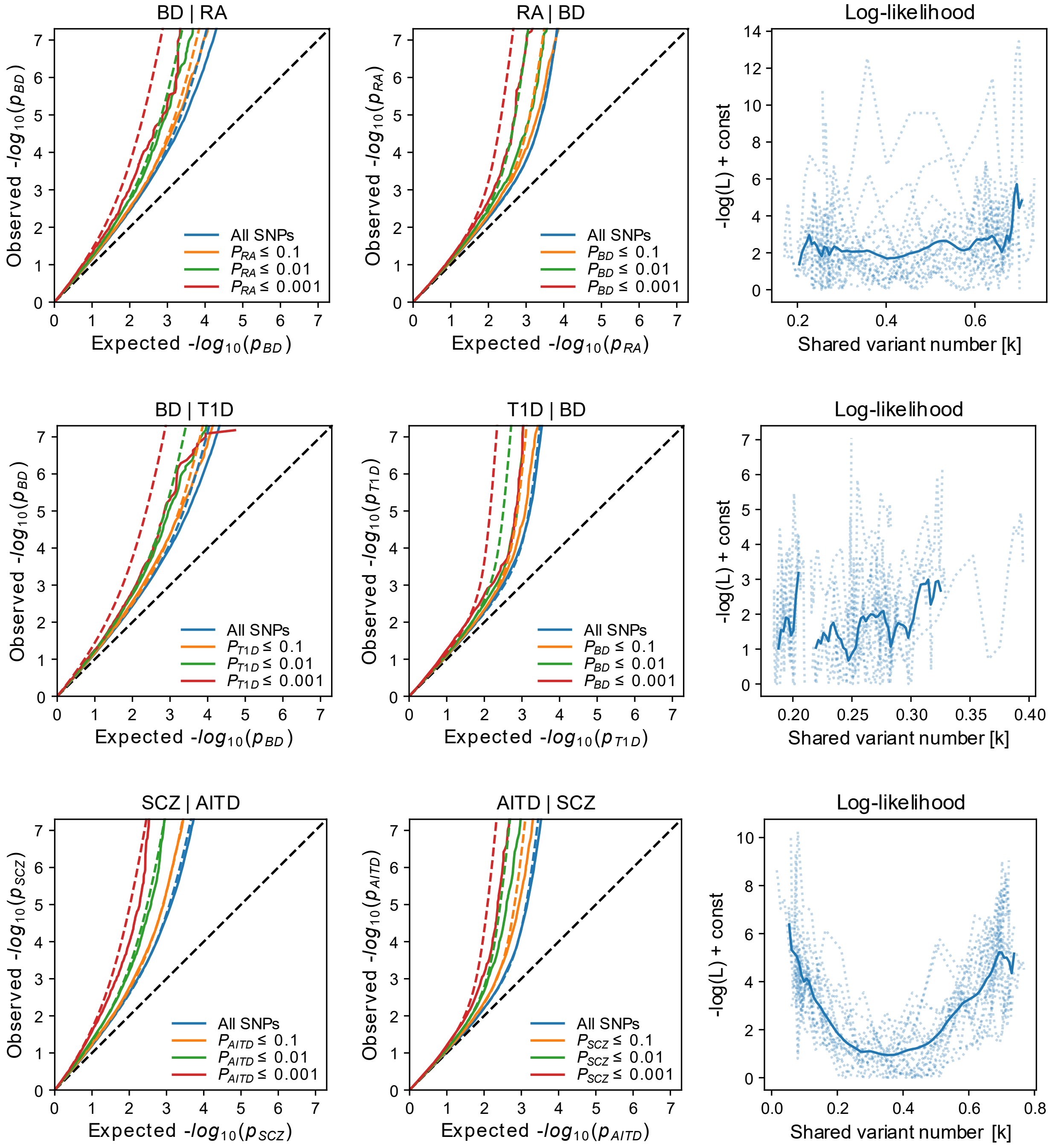
BD | RA
RA | BD
Log-likelihood
BD | T1D
T1D | BD
Log-likelihood
SCZ | AITD
AITD | SCZ
Log-likelihood
Observed -log10(pBD)
Observed -log10(pRA)
Observed -log10(pT1D)
Observed -log10(pSCZ)
Observed -log10(pAITD)
Expected -log10(pBD)
Expected -log10(pRA)
Expected -log10(pT1D)
Expected -log10(pSCZ)
Expected -log10(pAITD)
-log(L) + const
Shared variant number [k]
All SNPs
PRA ≤ 0.1
PRA ≤ 0.01
PRA ≤ 0.001
PBD ≤ 0.1
PBD ≤ 0.01
PBD ≤ 0.001
PT1D ≤ 0.1
PT1D ≤ 0.01
PT1D ≤ 0.001
PAITD ≤ 0.1
PAITD ≤ 0.01
PAITD ≤ 0.001
PSCZ ≤ 0.1
PSCZ ≤ 0.01
PSCZ ≤ 0.001

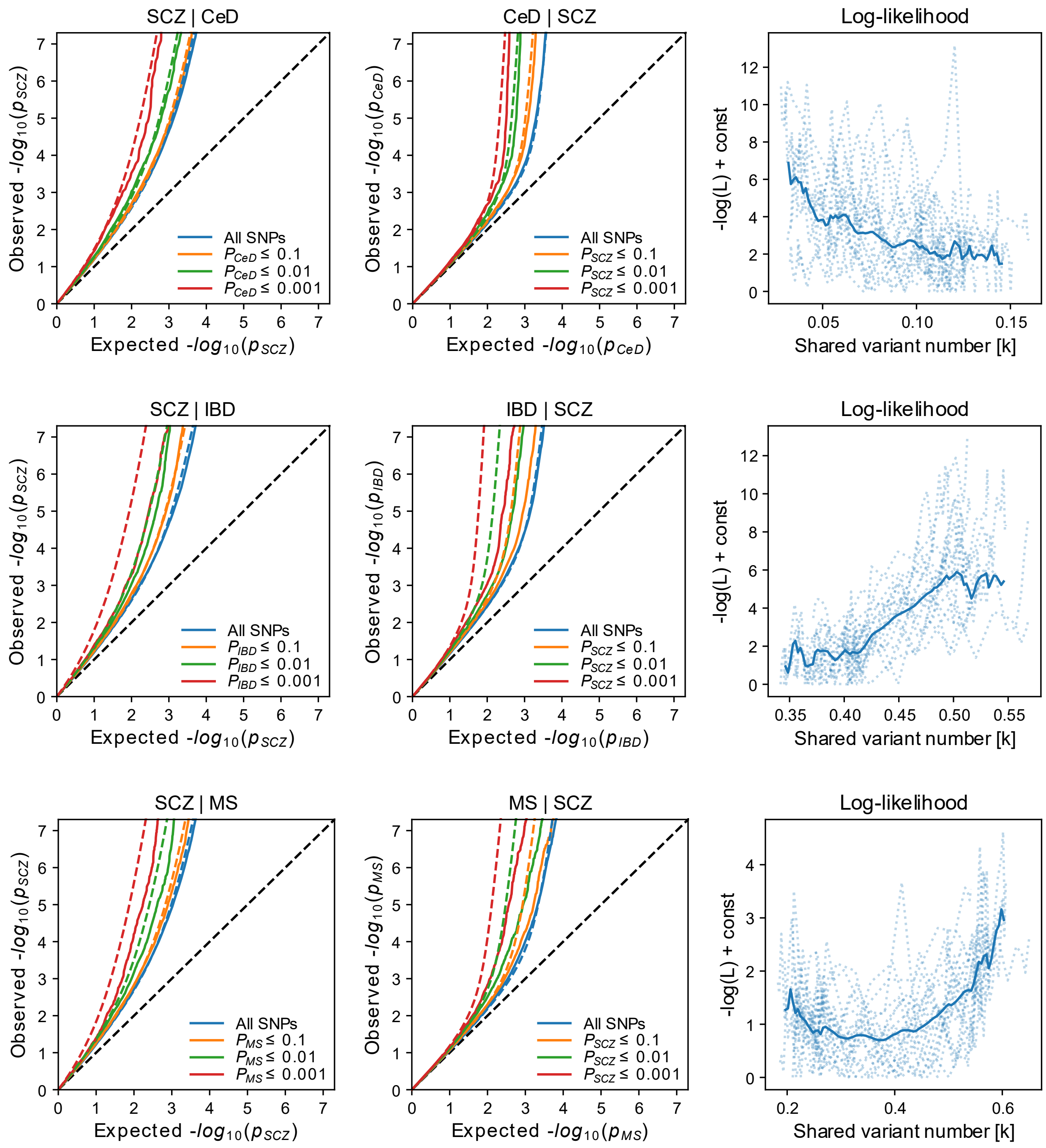
SCZ | CeD
CeD | SCZ
Log-likelihood
SCZ | IBD
IBD | SCZ
Log-likelihood
SCZ | MS
MS | SCZ
Log-likelihood
Observed -log10(pSCZ)
Observed -log10(pCeD)
Observed -log10(pIBD)
Observed -log10(pMS)
Expected -log10(pSCZ)
Expected -log10(pCeD)
Expected -log10(pIBD)
Expected -log10(pMS)
-log(L) + const
Shared variant number [k]
All SNPs
PCeD ≤ 0.1
PCeD ≤ 0.01
PCeD ≤ 0.001
PSCZ ≤ 0.1
PSCZ ≤ 0.01
PSCZ ≤ 0.001
PIBD ≤ 0.1
PIBD ≤ 0.01
PIBD ≤ 0.001
PMS ≤ 0.1
PMS ≤ 0.01
PMS ≤ 0.001

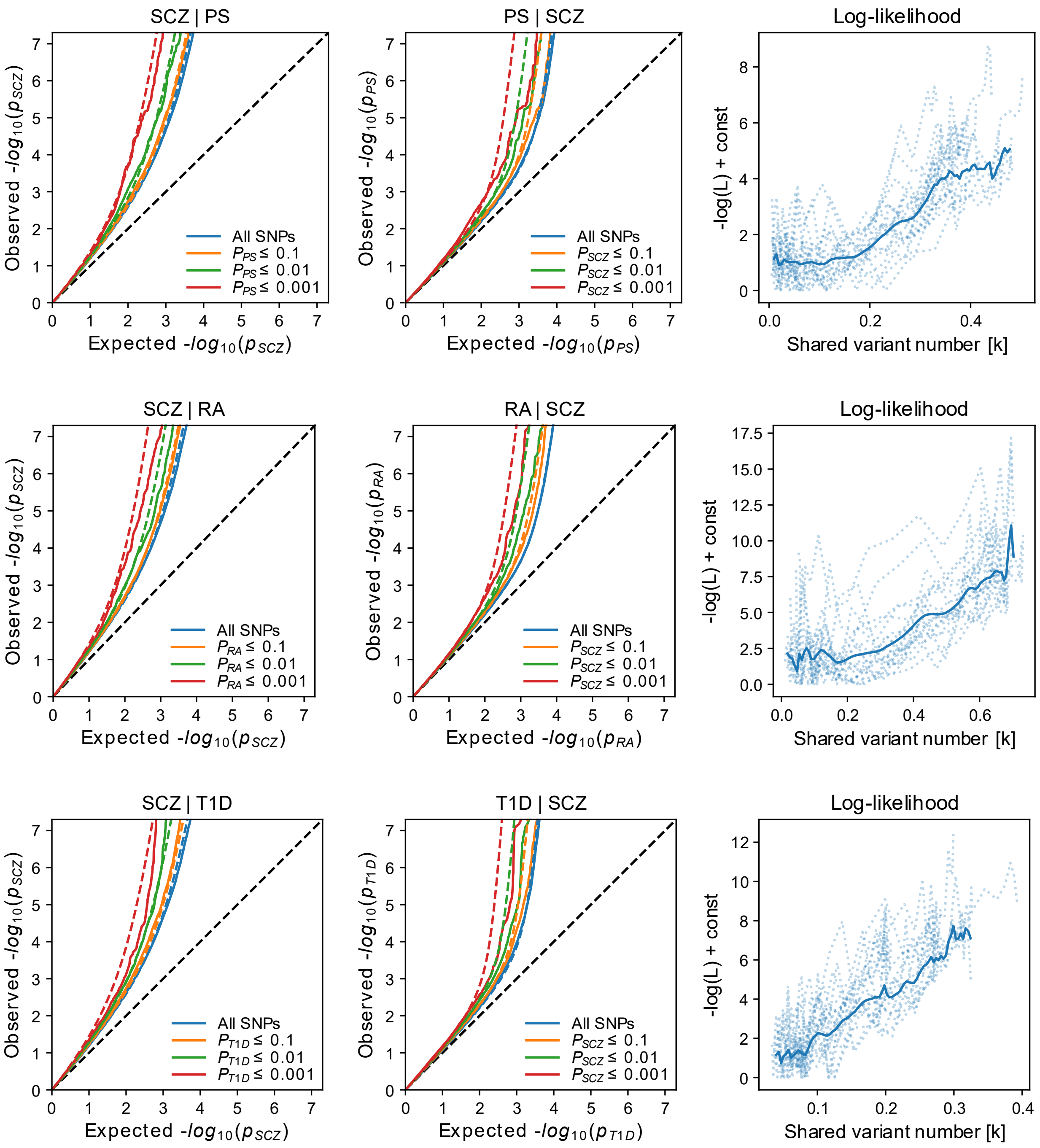

Conditional Q-Q plots display observed versus expected cross-trait enrichment, and log-likelihoods plots display the adjusted negative log-likelihood function against the number of shared causal variants, constrained to minimum and maximum possible overlap. Analyses were performed pairwise between the mental disorders major depression (MD), bipolar disorder (BD), and schizophrenia (SCZ), and the autoimmune diseases autoimmune thyroiditis (AITD), celiac disease (CeD), inflammatory bowel disease (IBD), multiple sclerosis (MS), psoriasis (PS), rheumatoid arthritis (RA), and type 1 diabetes (T1D).

**Supplementary Figure 3. GWAS power plots estimated by MiXeR**

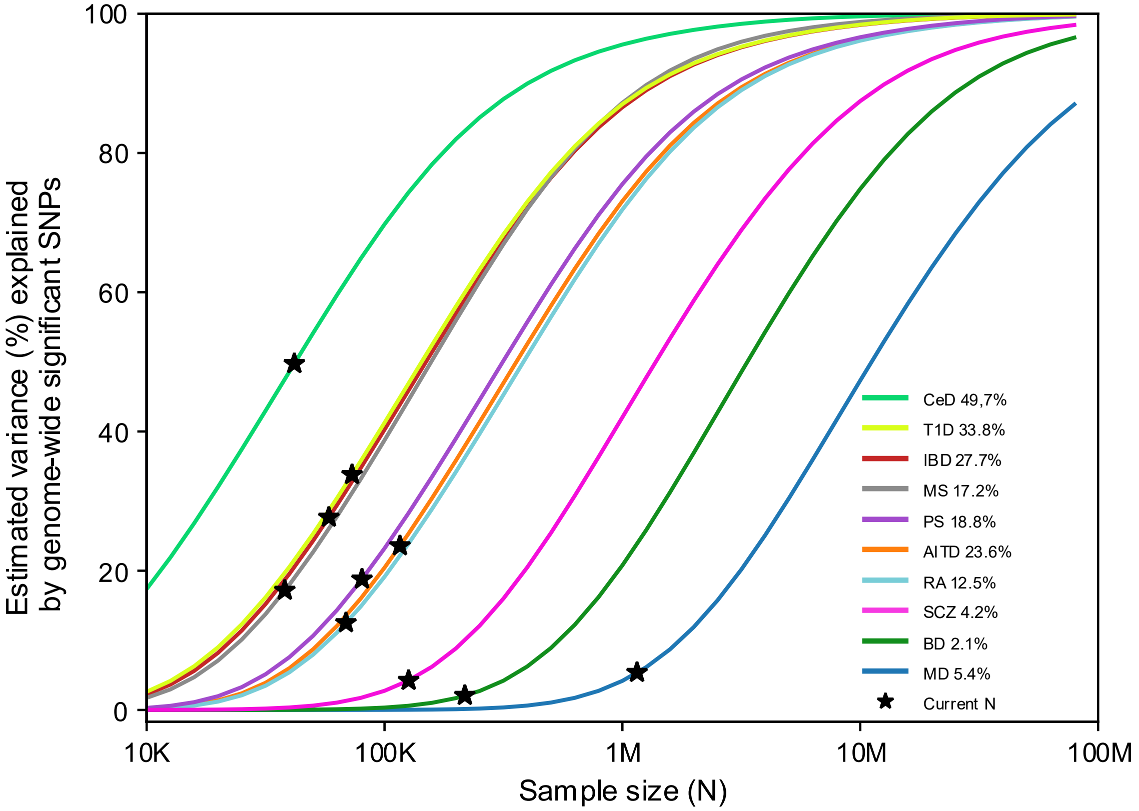

GWAS power plots display the proportion of SNP heritability explained by genome-wide significant SNPs as a function of effective sample size (N) for major depression (MD), bipolar disorder (BD), schizophrenia (SCZ), autoimmune thyroiditis (AITD), celiac disease (CeD), inflammatory bowel disease (IBD), multiple sclerosis (MS), psoriasis (PS), rheumatoid arthritis (RA), and type 1 diabetes (T1D). Asterisks indicate the current effective sample sizes of the respective GWASs.

**Supplementary Figure 4. Loci identified by Local Analysis of [co]Variant Association (LAVA)**

**a**

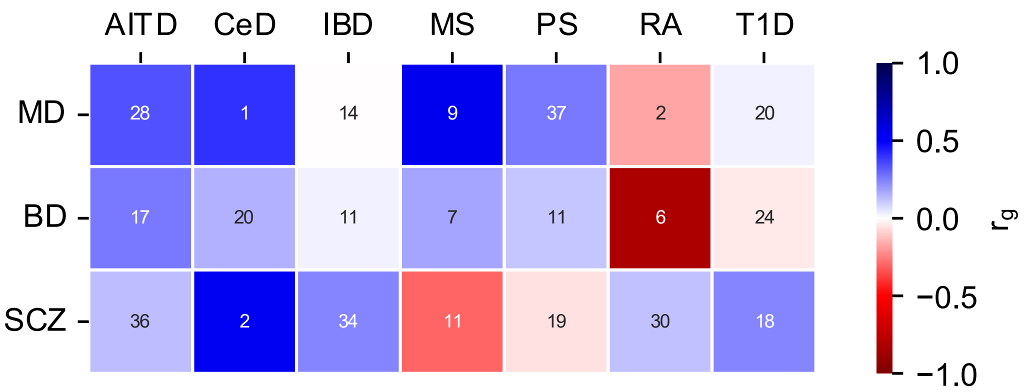

**b**

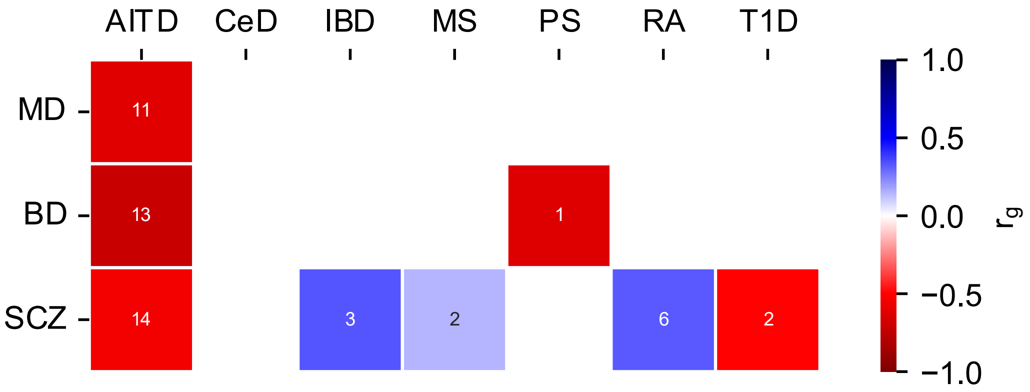

Number of genetic loci with significant local genetic correlations detected by LAVA outside the extended MHC region (**a**) and within the extended MHC region (**b**), between the mental disorders major depression (MD), bipolar disorder (BD), and schizophrenia (SCZ), and the autoimmune diseases autoimmune thyroiditis (AITD), celiac disease (CeD), inflammatory bowel disease (IBD), multiple sclerosis (MS), psoriasis (PS), rheumatoid arthritis (RA), and type 1 diabetes (T1D). Colors represent the mean correlation values ($r_g$s) within the significant loci.

**Supplementary Figure 5. Q-Q plots from conjFDR analyses**

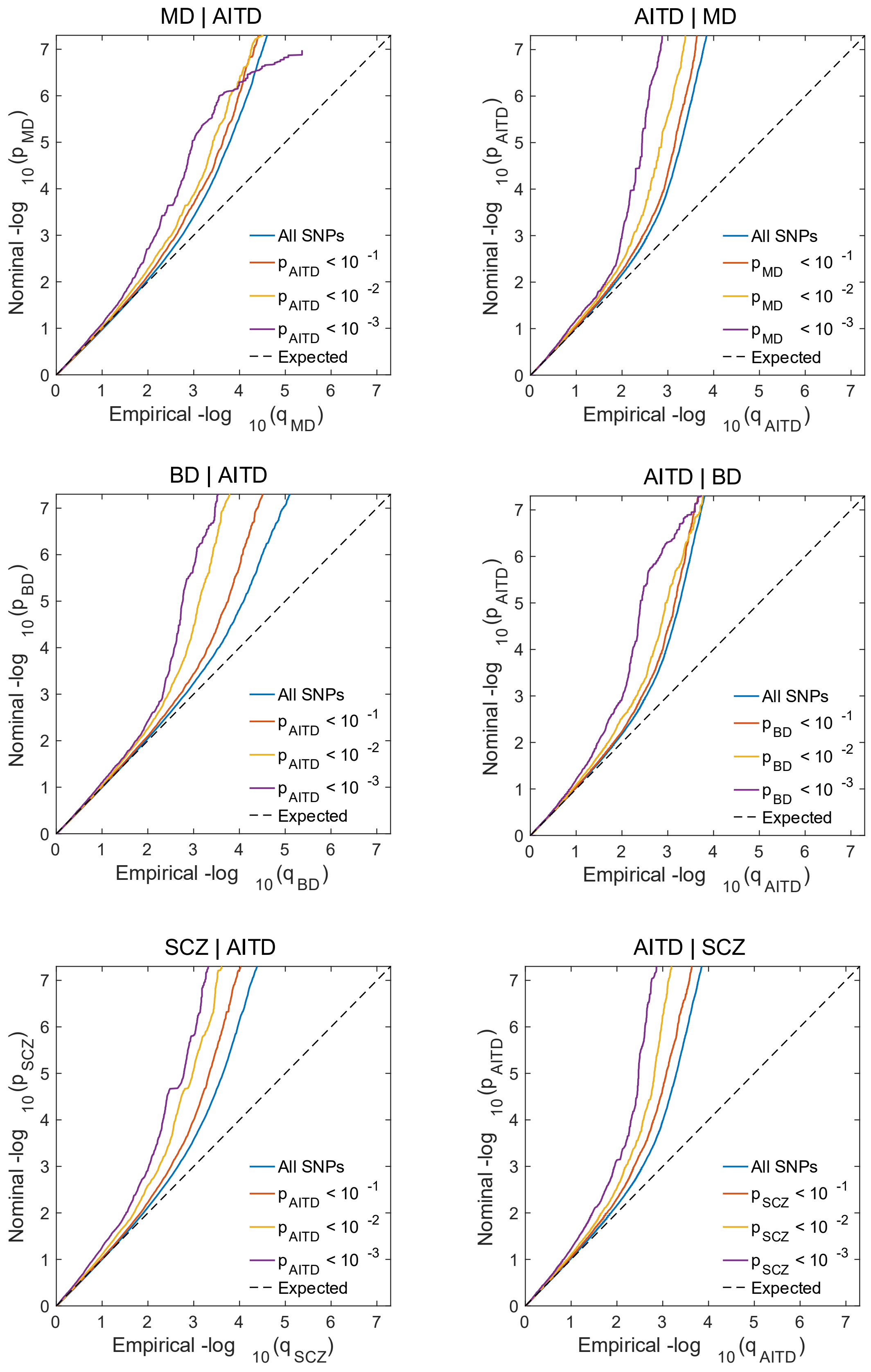

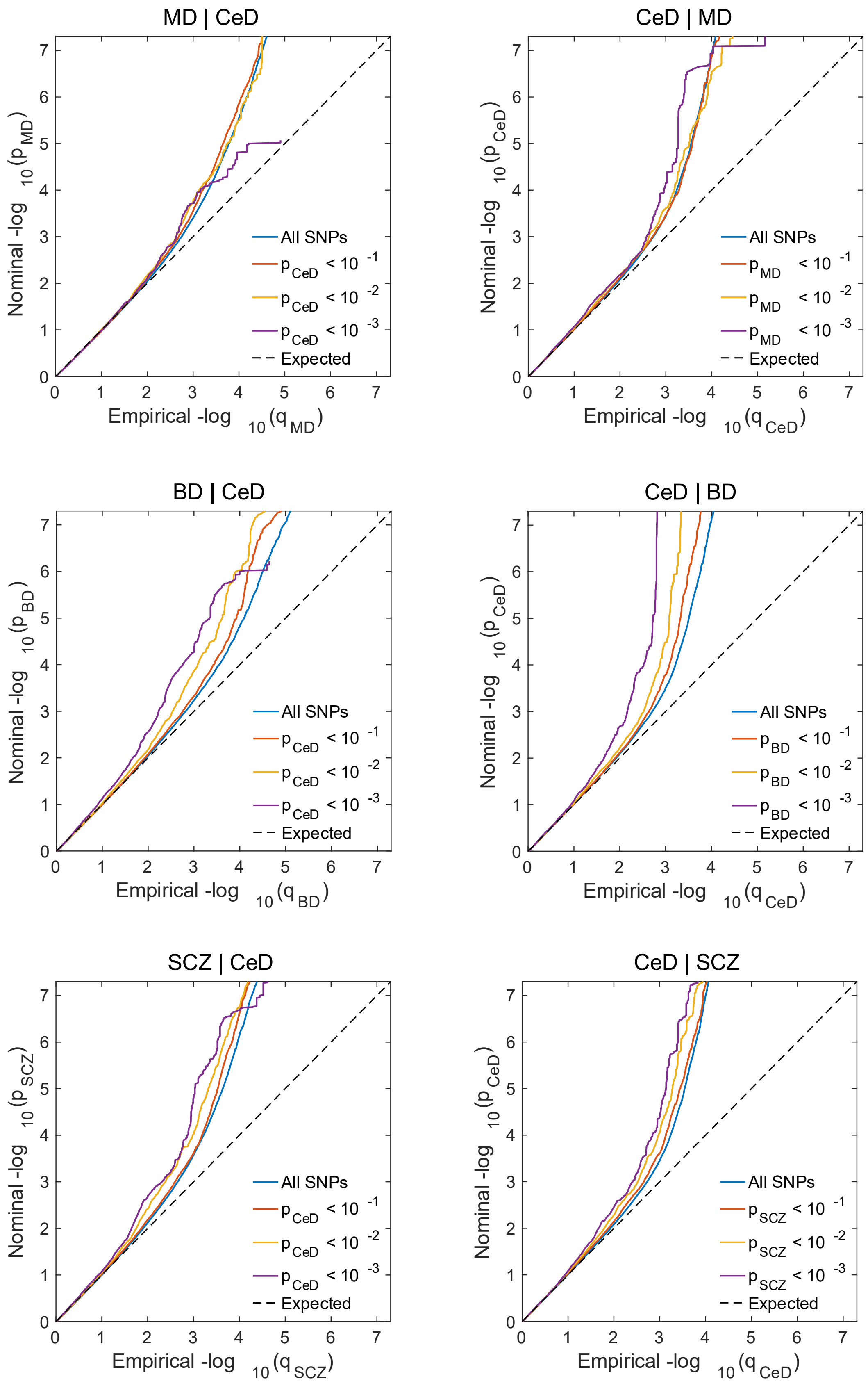
MD | CeD
CeD | MD
BD | CeD
CeD | BD
SCZ | CeD
CeD | SCZ
Nominal -log 10 (p MD)
Nominal -log 10 (p CeD)
Nominal -log 10 (p BD)
Nominal -log 10 (p SCZ)
Empirical -log 10 (q MD)
Empirical -log 10 (q CeD)
Empirical -log 10 (q BD)
Empirical -log 10 (q SCZ)
All SNPs
p CeD < 10 -1
p CeD < 10 -2
p CeD < 10 -3
p MD < 10 -1
p MD < 10 -2
p MD < 10 -3
p BD < 10 -1
p BD < 10 -2
p BD < 10 -3
p SCZ < 10 -1
p SCZ < 10 -2
p SCZ < 10 -3
Expected

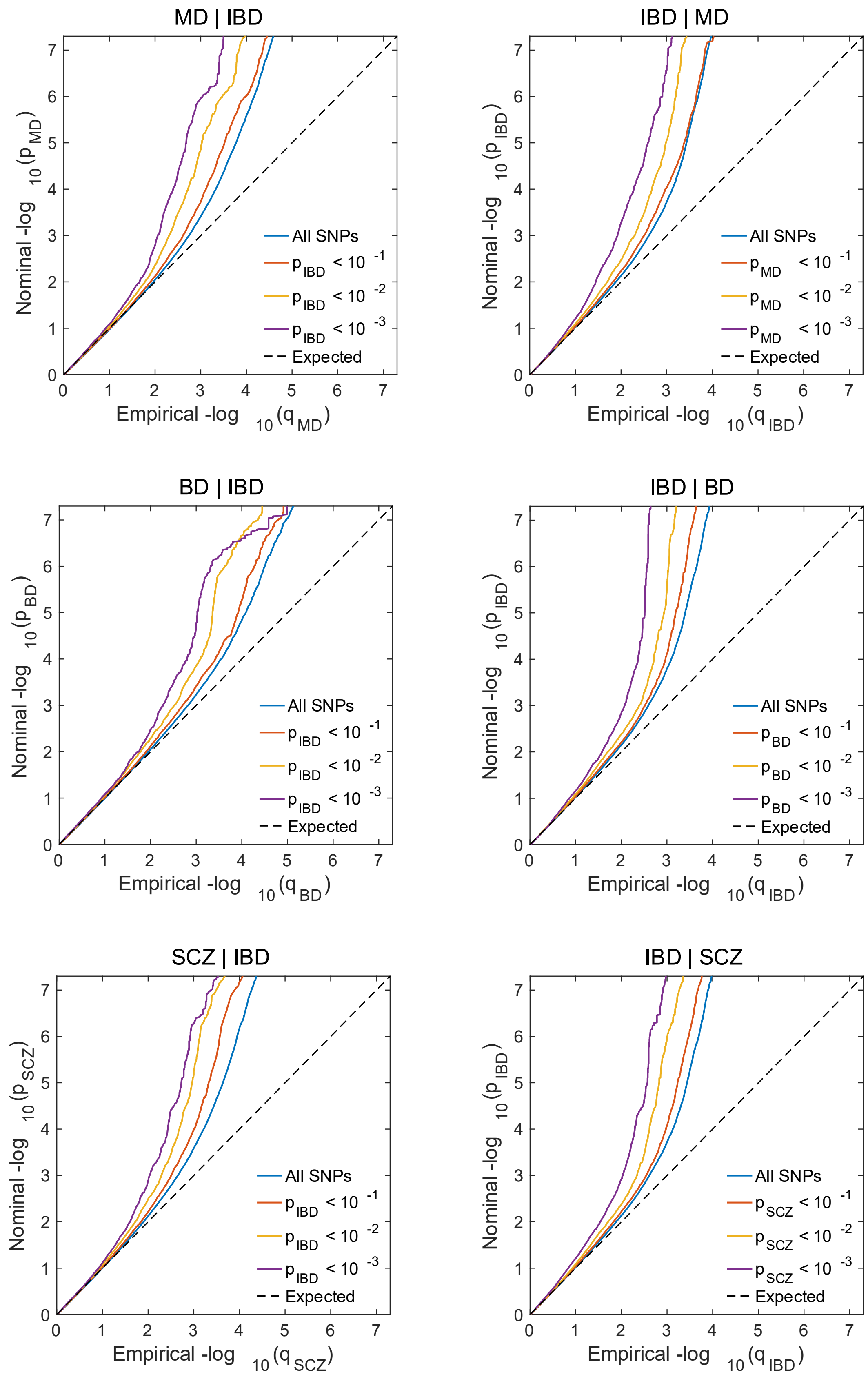
MD | IBD
Nominal -log 10(p MD)
Empirical -log 10(q MD)
All SNPs
p IBD < 10 -1
p IBD < 10 -2
p IBD < 10 -3
Expected
IBD | MD
Nominal -log 10(p IBD)
Empirical -log 10(q IBD)
All SNPs
p MD < 10 -1
p MD < 10 -2
p MD < 10 -3
Expected
BD | IBD
Nominal -log 10(p BD)
Empirical -log 10(q BD)
All SNPs
p IBD < 10 -1
p IBD < 10 -2
p IBD < 10 -3
Expected
IBD | BD
Nominal -log 10(p IBD)
Empirical -log 10(q IBD)
All SNPs
p BD < 10 -1
p BD < 10 -2
p BD < 10 -3
Expected
SCZ | IBD
Nominal -log 10(p SCZ)
Empirical -log 10(q SCZ)
All SNPs
p IBD < 10 -1
p IBD < 10 -2
p IBD < 10 -3
Expected
IBD | SCZ
Nominal -log 10(p IBD)
Empirical -log 10(q IBD)
All SNPs
p SCZ < 10 -1
p SCZ < 10 -2
p SCZ < 10 -3
Expected

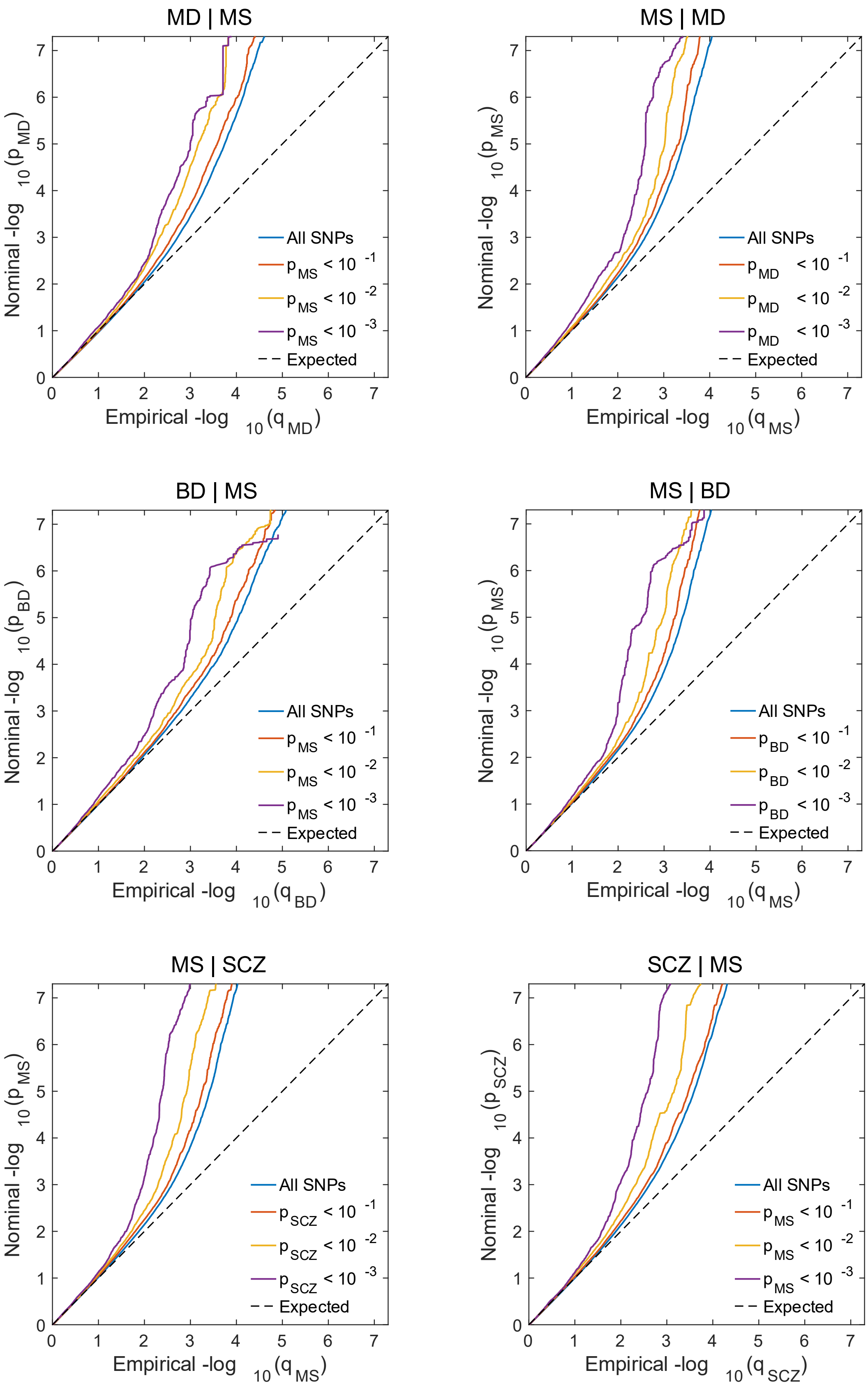
MD | MS
Nominal -log 10(p MD)
Empirical -log 10(q MD)
All SNPs
p MS < 10 -1
p MS < 10 -2
p MS < 10 -3
Expected
MS | MD
Nominal -log 10(p MS)
Empirical -log 10(q MS)
All SNPs
p MD < 10 -1
p MD < 10 -2
p MD < 10 -3
Expected
BD | MS
Nominal -log 10(p BD)
Empirical -log 10(q BD)
All SNPs
p MS < 10 -1
p MS < 10 -2
p MS < 10 -3
Expected
MS | BD
Nominal -log 10(p MS)
Empirical -log 10(q MS)
All SNPs
p BD < 10 -1
p BD < 10 -2
p BD < 10 -3
Expected
MS | SCZ
Nominal -log 10(p MS)
Empirical -log 10(q MS)
All SNPs
p SCZ < 10 -1
p SCZ < 10 -2
p SCZ < 10 -3
Expected
SCZ | MS
Nominal -log 10(p SCZ)
Empirical -log 10(q SCZ)
All SNPs
p MS < 10 -1
p MS < 10 -2
p MS < 10 -3
Expected

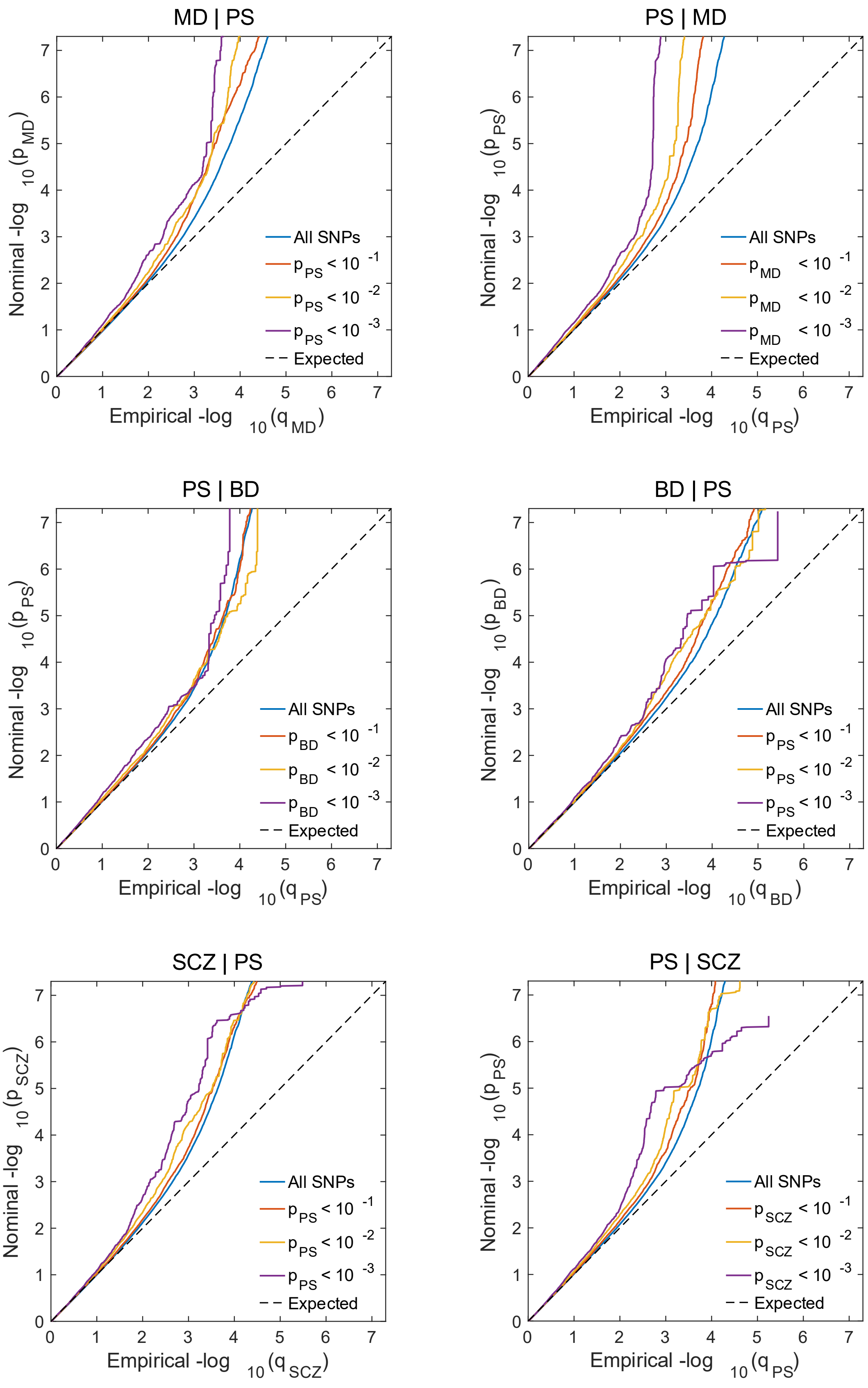
MD | PS
PS | MD
PS | BD
BD | PS
SCZ | PS
PS | SCZ
Nominal -log 10(p MD)
Nominal -log 10(p PS)
Nominal -log 10(p BD)
Nominal -log 10(p SCZ)
Empirical -log 10(q MD)
Empirical -log 10(q PS)
Empirical -log 10(q BD)
Empirical -log 10(q SCZ)
All SNPs
p PS < 10 -1
p PS < 10 -2
p PS < 10 -3
p MD < 10 -1
p MD < 10 -2
p MD < 10 -3
p BD < 10 -1
p BD < 10 -2
p BD < 10 -3
p SCZ < 10 -1
p SCZ < 10 -2
p SCZ < 10 -3
Expected

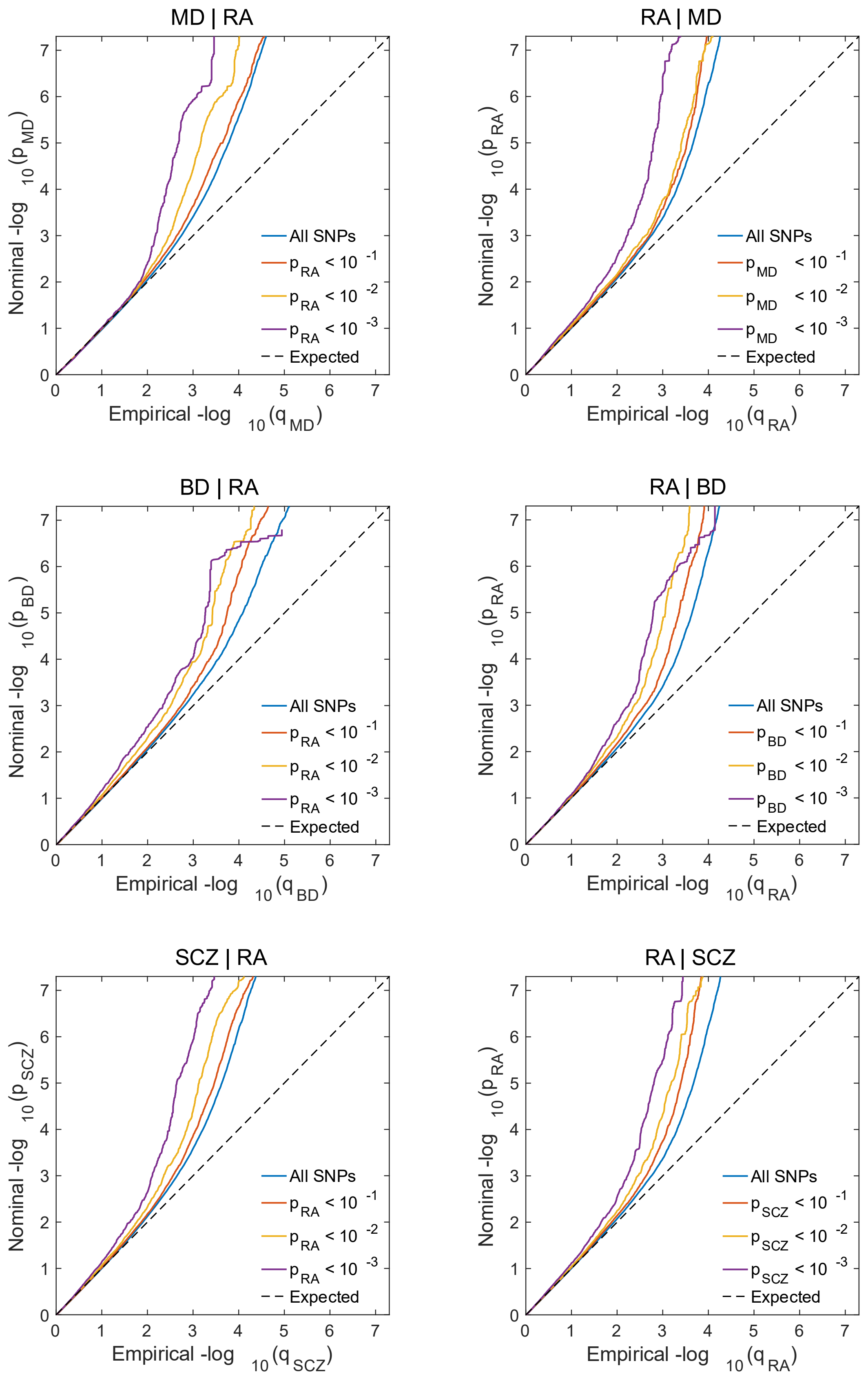
MD | RA
RA | MD
BD | RA
RA | BD
SCZ | RA
RA | SCZ
Nominal -log $_{10}$($p_{MD}$)
Nominal -log $_{10}$($p_{RA}$)
Nominal -log $_{10}$($p_{BD}$)
Nominal -log $_{10}$($p_{SCZ}$)
Empirical -log $_{10}$($q_{MD}$)
Empirical -log $_{10}$($q_{RA}$)
Empirical -log $_{10}$($q_{BD}$)
Empirical -log $_{10}$($q_{SCZ}$)
All SNPs
$p_{RA} < 10^{-1}$
$p_{RA} < 10^{-2}$
$p_{RA} < 10^{-3}$
$p_{MD} < 10^{-1}$
$p_{MD} < 10^{-2}$
$p_{MD} < 10^{-3}$
$p_{BD} < 10^{-1}$
$p_{BD} < 10^{-2}$
$p_{BD} < 10^{-3}$
$p_{SCZ} < 10^{-1}$
$p_{SCZ} < 10^{-2}$
$p_{SCZ} < 10^{-3}$
Expected

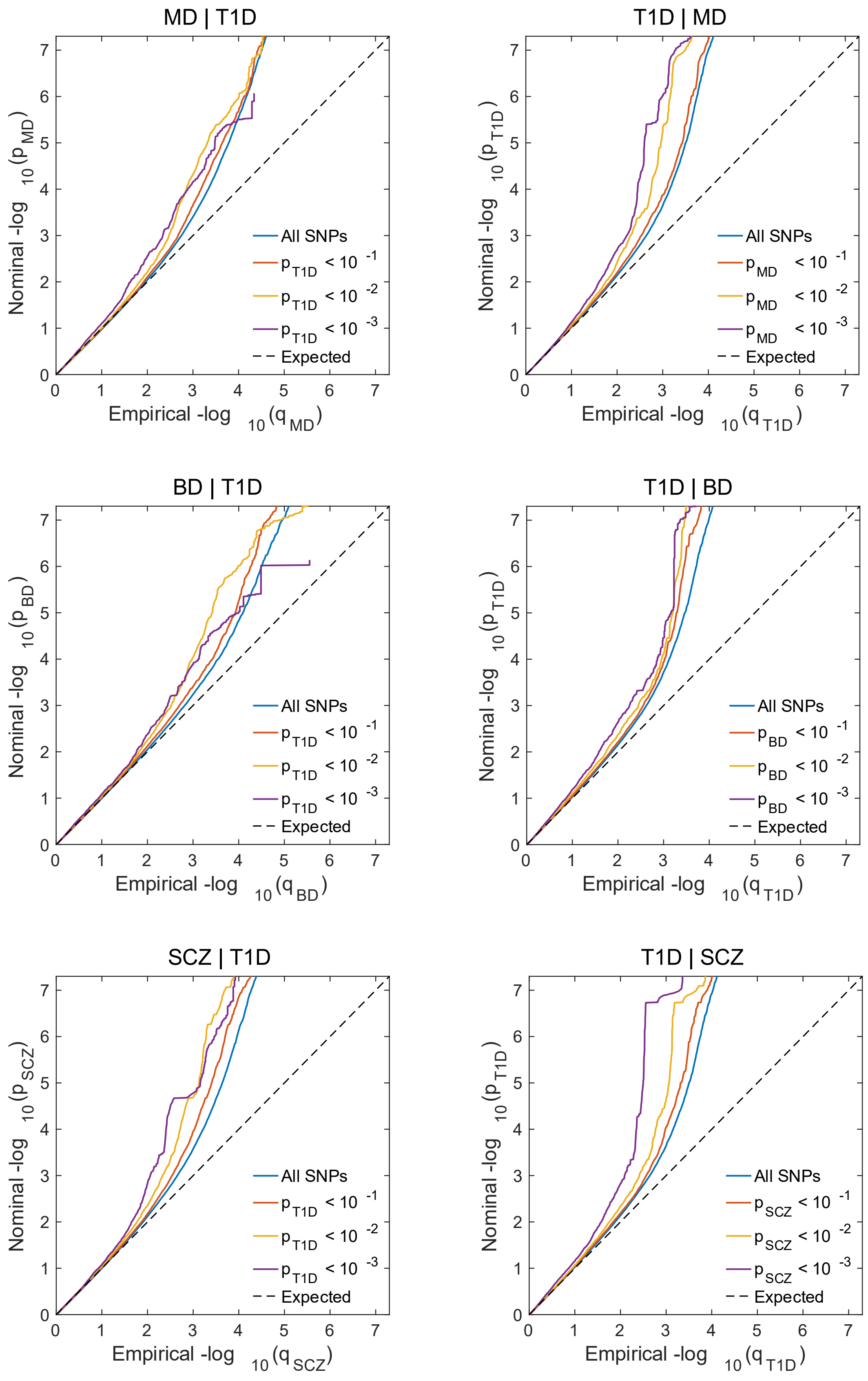
MD | T1D
Nominal -log 10(p MD)
Empirical -log 10(q MD)
All SNPs
p T1D < 10 -1
p T1D < 10 -2
p T1D < 10 -3
Expected
T1D | MD
Nominal -log 10(p T1D)
Empirical -log 10(q T1D)
All SNPs
p MD < 10 -1
p MD < 10 -2
p MD < 10 -3
Expected
BD | T1D
Nominal -log 10(p BD)
Empirical -log 10(q BD)
All SNPs
p T1D < 10 -1
p T1D < 10 -2
p T1D < 10 -3
Expected
T1D | BD
Nominal -log 10(p T1D)
Empirical -log 10(q T1D)
All SNPs
p BD < 10 -1
p BD < 10 -2
p BD < 10 -3
Expected
SCZ | T1D
Nominal -log 10(p SCZ)
Empirical -log 10(q SCZ)
All SNPs
p T1D < 10 -1
p T1D < 10 -2
p T1D < 10 -3
Expected
T1D | SCZ
Nominal -log 10(p T1D)
Empirical -log 10(q T1D)
All SNPs
p SCZ < 10 -1
p SCZ < 10 -2
p SCZ < 10 -3
Expected

Conditional Q-Q plots illustrate cross-trait enrichment between the mental disorders major depression (MD), bipolar disorder (BD), and schizophrenia (SCZ), and the autoimmune diseases autoimmune thyroiditis (AITD), celiac disease (CeD), inflammatory bowel disease (IBD), multiple sclerosis (MS), psoriasis (PS), rheumatoid arthritis (RA), and type 1 diabetes (T1D). The x-axis represents empirical - $\log_{10}$ *P* values, while the y-axis represents nominal - $\log_{10}$ *P* values. The dotted black line shows the expected values for all SNPs under the null hypothesis of no polygenicity. The blue line shows the distribution of *P* values for all SNPs, while the red, yellow and purple lines represent subset of SNPs associated with the secondary trait at increasing *P* values.

**Supplementary Figure 6. Tissue-specific expression from FUMA gene set analyses**

**a**

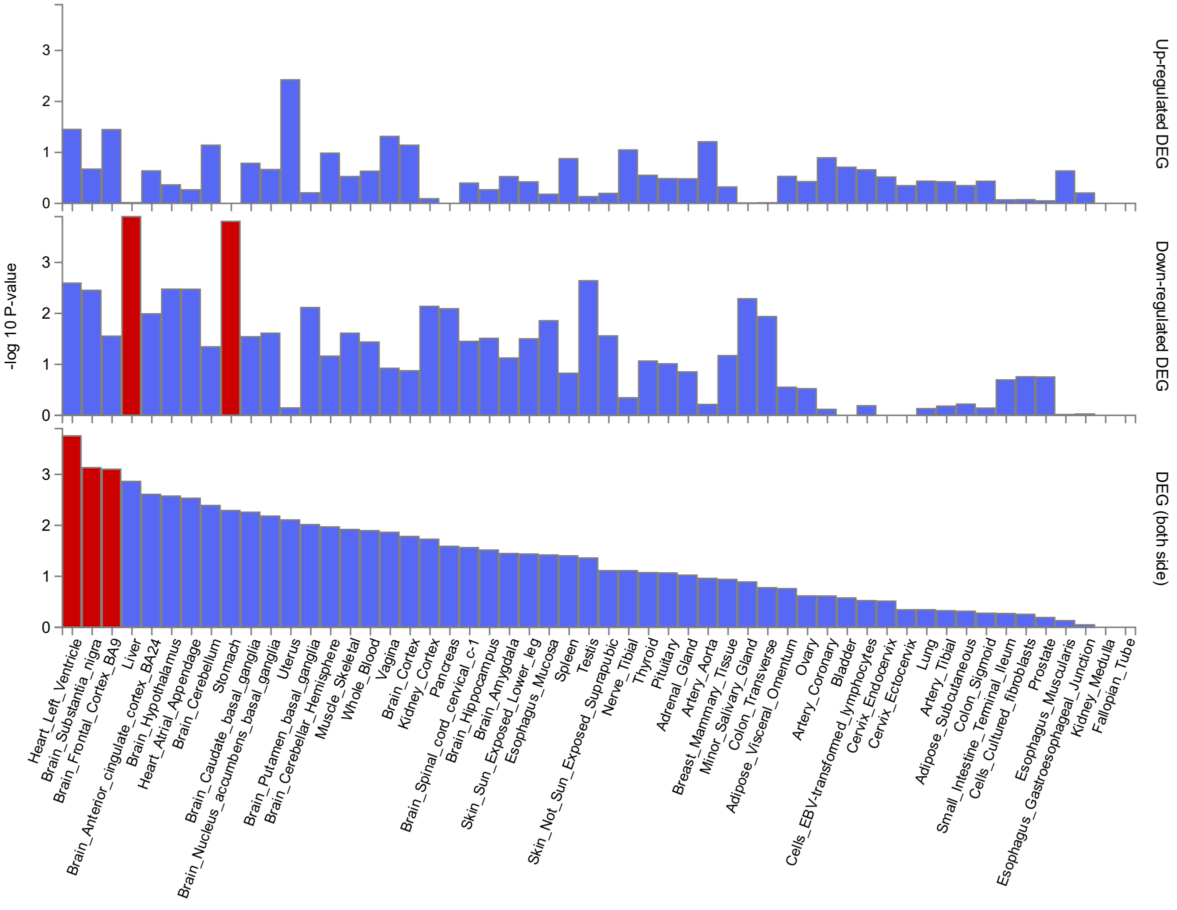

**b**

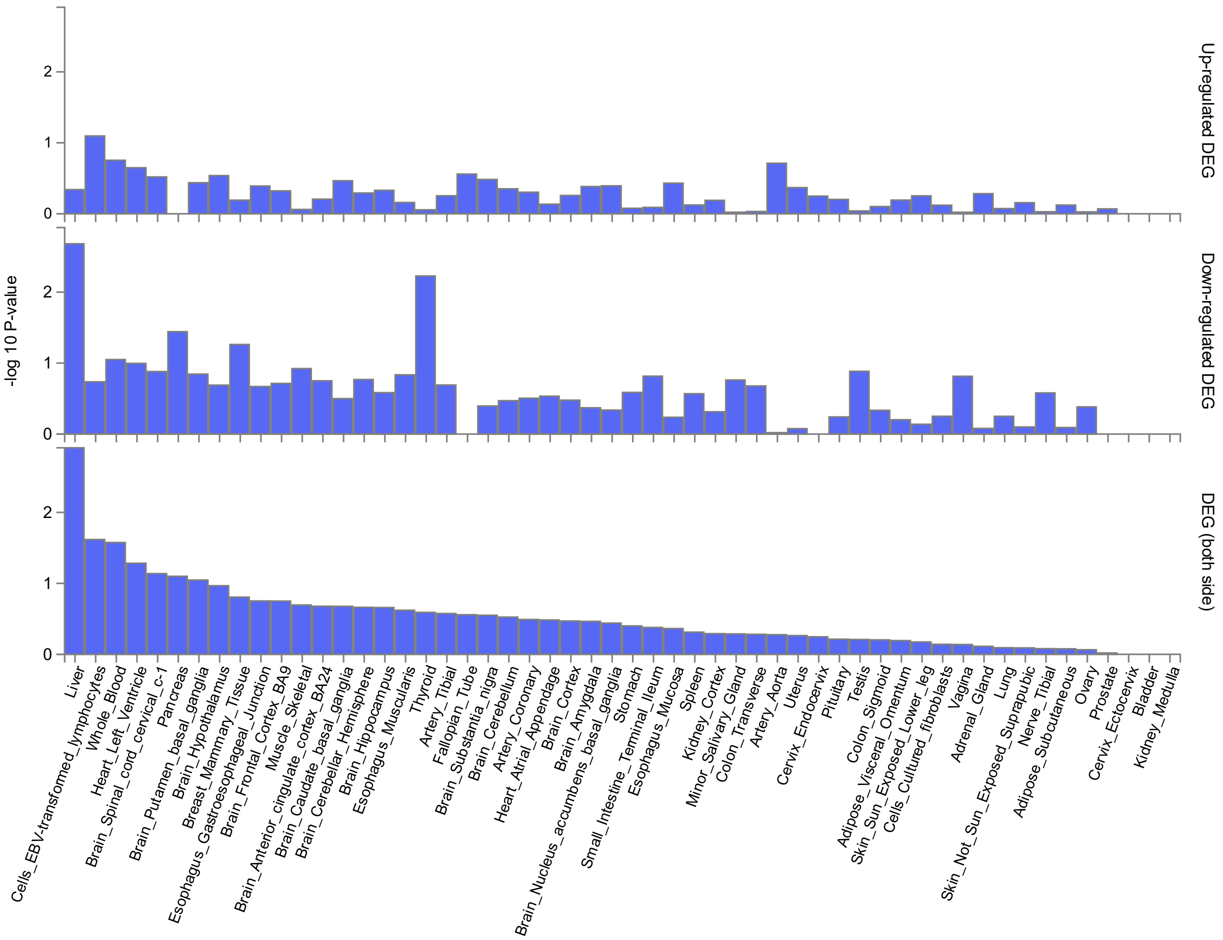

Up-regulated DEG
Down-regulated DEG
DEG (both side)
-log 10 P-value
0
1
2
Liver
Cells_EBV-transformed_lymphocytes
Whole_Blood
Heart_Left_Ventricle
Brain_Spinal_cord_cervical_c-1
Pancreas
Brain_Putamen_basal_ganglia
Brain_Hypothalamus
Breast_Mammary_Tissue
Esophagus_Gastroesophageal_Junction
Brain_Frontal_Cortex_BA9
Muscle_Skeletal
Brain_Anterior_cingulate_cortex_BA24
Brain_Caudate_basal_ganglia
Brain_Cerebellar_Hemisphere
Brain_Hippocampus
Esophagus_Muscularis
Thyroid
Artery_Tibial
Fallopian_Tube
Brain_Substantia_nigra
Brain_Cerebellum
Artery_Coronary
Heart_Atrial_Appendage
Brain_Cortex
Brain_Amygdala
Brain_Nucleus_accumbens_basal_ganglia
Stomach
Small_Intestine_Terminal_Ileum
Esophagus_Mucosa
Spleen
Kidney_Cortex
Minor_Salivary_Gland
Colon_Transverse
Artery_Aorta
Uterus
Cervix_Endocervix
Pituitary
Testis
Colon_Sigmoid
Adipose_Visceral_Omentum
Skin_Sun_Exposed_Lower_leg
Cells_Cultured_fibroblasts
Vagina
Adrenal_Gland
Lung
Skin_Not_Sun_Exposed_Suprapubic
Nerve_Tibial
Adipose_Subcutaneous
Ovary
Prostate
Cervix_Ectocervix
Bladder
Kidney_Medulla

**c**

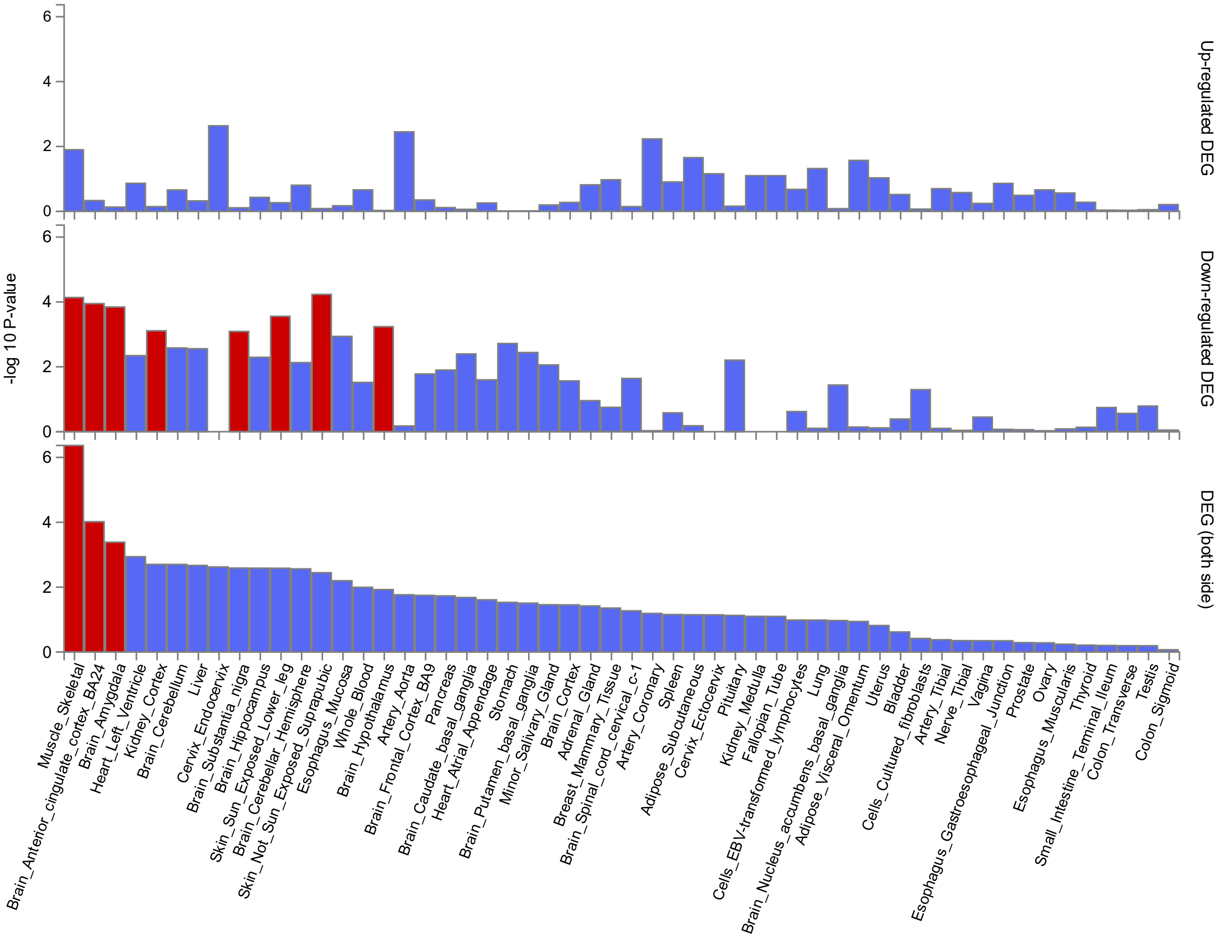

Column charts illustrate the tissue-specific regulation of genes identified by conjFDR and Open Targets, shared between major depression (**a**), bipolar disorder (**b**), and schizophrenia (**c**), and at least one of the autoimmune diseases: autoimmune thyroiditis, celiac disease, inflammatory bowel disease, multiple sclerosis, psoriasis, rheumatoid arthritis, and type 1 diabetes. Gene set analysis was performed using FUMA based on gene expression data from 54 tissue types available in GTEx v8. The MHC region was excluded from the analyses. The charts indicate whether the gene set is upregulated, downregulated or differentially regulated (upregulated and downregulated) across tissues. Statistically significant tissues after Bonferroni correction are colored red. The y-axis shows $-\log_{10}$ *P* values, and the x-axis shows 53 different tissue types.

### Phenotype definitions

Main analysis

*Major depression (MD)*

Data on MD were collected from cohorts based in Europe, Oceania, and North America (1). The definition of the MD phenotype varied across individual studies. In some cohorts, diagnosis was based on self-reporting, while others relied on electronic health records, with diagnoses determined by medical professionals based on International Classification of Diseases (ICD) or Diagnostic and Statistical Manual of Mental Disorders (DSM) criteria.

*Bipolar disorder (BD)*

Data on BD were obtained from cohorts based in Europe, Oceania and North America (2). The diagnosis was largely established based on consensus criteria from either ICD or DSM, derived through assessments by trained interviewers or through medical record review. Some cases were based on individuals self-reporting having received the diagnosis from a medical professional.

*Schizophrenia (SCZ)*

SCZ data were also collected from cohorts based in Europe, Oceania and North America (3). Diagnoses were confirmed by specialists or trained researchers based on ICD or DSM consensus criteria.

*Autoimmune thyroiditis (AITD)*

Data on AITD were gathered from deCODE Genetics, a nation-wide research program in Iceland utilizing hospital records for diagnosis, and the UK Biobank, a large prospective cohort study in the United Kingdom relying on electronic health records from both hospital data as well as primary care for diagnosis (4). Individuals with AITD were identified using a broad disease definition, including Graves' disease, Hashimoto's thyroiditis, and unspecified hypothyroidism with the exclusion of other known causes of thyroid dysfunction, such as thyroid cancer or drug-induced hypothyroidism.

*Celiac disease (CeD)*

We conducted our own genome-wide association study (GWAS) on CeD from cohorts from Norway, Finland, and the UK. In data from the Norwegian Mother, Father and Child Cohort Study (MoBa) and the Hordaland Health Study (HUSK), we confirmed the diagnosis based on electronic health records according to ICD diagnostic criteria. UK Biobank relied on electronic health records from both hospital data and primary care for diagnosis. FinnGen also relied on electronic health records, with the diagnosis defined according to ICD diagnostic criteria. MoBa is a population-based pregnancy cohort study conducted by the Norwegian Institute of Public Health. Participants were recruited from all over Norway from 1999-2008. The women consented to participation in 41% of the pregnancies. The cohort includes approximately 114,500 children, 95,200 mothers and 75,200 fathers. MoBa is regulated by the Norwegian Health Registry Act. The FinnGen study is a large-scale genomics initiative that has analyzed over 500,000 Finnish biobank samples and correlated genetic variation with health data to understand disease mechanisms and predispositions. The project is a collaboration between research organizations and biobanks within Finland and international industry partners. The HUSK study was conducted by the University of Bergen in cooperation with the Norwegian Institute of Public Health.

*Inflammatory bowel disease (IBD)*

Data on IBD were gathered from cohorts in Europe, Oceania, and North America. Participants were diagnosed according to accepted endoscopic, histopathological, and radiological criteria (5).

*Multiple sclerosis (MS)*

Data on MS included cohorts from Europe, Oceania, and North America. MS cases were diagnosed by neurologists in accordance with standard criteria, though some cohorts relied on self-reporting (6).

*Psoriasis (PS)*

We performed our own GWAS on PS, collecting data from cohorts in Norway, Finland and the UK. PS cases were determined by electronic health records according to ICD diagnostic criteria in MoBa, while in the HUSK dataset, the diagnosis was determined utilizing the International Classification of Primary Care, 2nd edition (ICPC-2) diagnostic system.

*Rheumatoid arthritis (RA)*

Data on RA were derived from cohorts from Europe and North America. All RA cases fulfilled the 1987 American College of Rheumatology (ACR) diagnostic criteria or the 2010 ACR/European League Against Rheumatism diagnostic criteria, or were diagnosed by a professional rheumatologist (7).

*Type 1 diabetes (T1D)*

Data on T1D were collected from European and North American cohorts. Cases were defined based on accepted diagnostic criteria (8).

Sensitivity analysis

In sensitivity linkage disequilibrium score regression (LDSC) analyses, autoimmune and MD cases in the UK Biobank (application number 27412) were determined according to ICD-diagnostic criteria utilizing electronic health records.

### Additional information on statistical analyses and ethical approvals

Data analysis

In our study, all GWAS summary statistics were aligned to dbSNP using the Cleansumstats Pipeline v.1.7.0 (https://doi.org/10.5281/ZENODO.7441266), removing indels, ambiguous SNPs (A/T and C/G), and missing rs-IDs (9,10).

In-house GWASs on CeD and PS

We performed our own GWASs using METAL (https://github.com/statgen/METAL) on CeD and PS to ensure sufficient power for subsequent analyses. We used publicly available data from UK Biobank and FinnGen, and data from MoBa and HUSK obtained through collaborative efforts (11–14). The GWASs on MoBa and HUSK were performed using Regenie v.3.5 (https://github.com/rgcgithub/regenie/releases/tag/v3.5) (15). Variants with a minor allele count below 20 were excluded. Covariates for HUSK were sex, year of birth (standardized), genotyping batch (two batches, encoded with one dummy variable), and 40 genetic principal components (PCs). Covariates for MoBa were sex, year of birth (standardized), genotyping batch (26 batches, encoded with 25 dummy variables), and 10 genetic PCs. Quality control in MoBa is described in https://www.biorxiv.org/content/10.1101/2022.06.23.496289v4 (16). We acquired UK Biobank data under application number 27412.

*Detailed quality control (QC) in HUSK*

HUSK participants were genotyped at deCODE Genetics (https://www.decode.com/) in two batches (10,083 and 25,432 individuals in batch 1 and 2 respectively) using customized Illumina GSA v3 array (687316 genotyped variants). Before phasing and imputation genotyping data were QCed separately for each batch using plink2. QC steps for variants included removal of indels, non-ATGC, strand ambiguous, poorly genotyped (plink2's option: --geno 0.05), non-autosomal and rare (--maf 0.05) variants as well as variants with extreme deviation from Hardy-Weinberg equilibrium (--hwe 1E-50 midp keep-fewhet). Chromosomal strand and allele order for the remaining variants were aligned based on the HGDP + 1KG reference panel from gnomAD (https://gnomad.broadinstitute.org/downloads#v3-hgdp-1kg) (17), and variants with alleles that were not possible to align to the reference as well as variants with MAF deviating from the reference by more than 30% were removed. Resulting genotypes from two batches were merged, variants with high missingness (--geno 0.02) and individuals with poor genotyping rate (--mind 0.05) were removed from the merged data. The combined dataset was then phased and imputed with Beagle 5.4 (version 27May24.118) using the HGDP + 1KG reference panel. Imputed data were further QCed by removing variants with low imputation quality (Beagle's DR2<0.5) and variants with minor allele frequency below 0.5% (--maf 0.005). The resulting dataset contained 10,531,075 variants and 35,476 individuals.

Description of the conjunctional false discovery rate (conjFDR) method

ConjFDR is an extension of the conditional FDR (condFDR) approach, and identifies genetic loci that affect two phenotypes by leveraging their cross-trait enrichment. In this process, the conditional FDR (condFDR) is first calculated for each SNP for a primary phenotype, where test statistics are reranked according to their association with a secondary phenotype. Then the second condFDR value is estimated by switching the primary and secondary phenotypes. After that, the maximum of two condFDR estimates defines the conjFDR for the SNP association with both phenotypes.

We utilized conditional Q-Q plots to assess cross-trait enrichment (18), displaying the negative $\log_{10}$-scaled *P* values for all SNPs affecting a primary phenotype, and for subsets of SNPs that are associated with a secondary phenotype at *P* values with increasing levels of significance. Evidence of genetic overlap is demonstrated if the curves reveal successive leftward deflection for subsets of SNPs with increasing significance in the secondary phenotype.

Locus definition in conjFDR analysis

Independent significant SNPs were identified as variants with a conjFDR <0.05 and in LD $r^2$ <0.6 relative to each other. A subset of these at approximately LD $r^2$ <0.1 of each other were chosen as lead SNPs. LD $r^2$ values were derived from the 1000 Genomes Project European-ancestry haplotype reference panel. We determined the border of each locus by identifying all candidate SNPs, which were defined as all SNPs with conjFDR <0.1 and in LD $r^2 \geq 0.6$ with a lead variant. If two loci were less than 250 kb apart, they were merged, with the most significant SNP designated as the lead SNP of the merged locus.

Functional annotation of lead SNPs implicated by conjFDR

Open Targets utilizes machine learning to link SNPs to genes based on genetic associations from a large aggregate of human GWASs and functional genomics data, including gene expression, chromatin interaction, protein abundance, and conformation data from a wide array of cell types and tissues. The likelihood that a lead SNP functionally implicates a gene is given as the variant to gene (V2G) score. For each lead SNP, we selected the gene with the highest V2G score. If two genes had identical scores, we examined the physically closest candidate SNPs. If a lead SNP could not be found in the database, we investigated the closest candidate SNP instead.

Ethical approvals

All participants provided informed consent. The included studies had ethical approvals from institutional review boards or equivalent committees. The Norwegian Institutional Review Board for the South-East Norway Region has determined that no additional approval is required for the use of anonymized group-level data (ref. 2011/1980). Access to individual level UK Biobank data was obtained under the Application Number 27412. The establishment of MoBa and initial data collection was based on a license from the Norwegian Data Protection Agency and an approval from The Regional Committees for Medical and Health Research Ethics. The MoBa cohort is currently regulated by the Norwegian Health Registry Act. Participants in FinnGen provided informed consent for biobank research on basis of the Finnish Biobank Act. Alternatively, separate research cohorts, collected before the Finnish Biobank Act came into effect (in September 2013) and the start of FinnGen (August 2017) were collected on the basis of study-specific consent and later transferred to the Finnish biobanks after approval by Fimea, the National Supervisory Authority for Welfare and Health. The current HUSK study was approved by the Regional Committee for Medical and Health Research Ethics of Western Norway.

### Supplementary results

Discoverability according to causal mixture model (MiXeR) analysis

We generated GWAS power plots visualizing the proportion of SNP heritability ($h^2_{SNP}$) explained by genome-wide significant SNPs as a function of effective sample size (Supplementary Figure 3). In line with their lower polygenicity but higher discoverability, a larger fraction of the common variant architecture for autoimmune diseases is captured when compared to mental disorders at comparable GWAS sample sizes. For example, genome-wide significant variants from the latest GWAS on BD only explain 2.3% of its $h^2_{SNP}$, compared to 49.7% for CeD variants, although the BD GWAS has a more than five times larger effective sample size.

**Supplementary references**

1. Adams MJ, Streit F, Meng X, Awasthi S, Adey BN, Choi KW, et al. Trans-ancestry genome-wide study of depression identifies 697 associations implicating cell types and pharmacotherapies. Cell. 2025 Feb 6;188(3):640-652.e9.

2. O'Connell KS, Koromina M, van der Veen T, Boltz T, David FS, Yang JMK, et al. Genomics yields biological and phenotypic insights into bipolar disorder. Nature. 2025 Mar;639(8056):968–75.

3. Trubetskoy V, Panagiotaropoulou G, Awasthi S, Braun A, Kraft J, Skarabis N, et al. Mapping genomic loci implicates genes and synaptic biology in schizophrenia. Nature. 2022 Apr;604(7906):502–8.

4. Saevarsdottir S, Olafsdottir TA, Ivarsdottir EV, Halldorsson GH, Gunnarsdottir K, Sigurdsson A, et al. FLT3 stop mutation increases FLT3 ligand level and risk of autoimmune thyroid disease. Nature. 2020 Aug;584(7822):619–23.

5. de Lange KM, Moutsianas L, Lee JC, Lamb CA, Luo Y, Kennedy NA, et al. Genome-wide association study implicates immune activation of multiple integrin genes in inflammatory bowel disease. Nat Genet. 2017 Feb;49(2):256–61.

6. International Multiple Sclerosis Genetics Consortium. Multiple sclerosis genomic map implicates peripheral immune cells and microglia in susceptibility. Science. 2019 Sep 27;365(6460):eaav7188.

7. Ishigaki K, Sakaue S, Terao C, Luo Y, Sonehara K, Yamaguchi K, et al. Multi-ancestry genome-wide association analyses identify novel genetic mechanisms in rheumatoid arthritis. Nat Genet. 2022 Nov;54(11):1640–51.

8. Chiou J, Geusz RJ, Okino ML, Han JY, Miller M, Melton R, et al. Interpreting type 1 diabetes risk with genetics and single-cell epigenomics. Nature. 2021 Jun;594(7863):398–402.

9. Wheeler DL, Barrett T, Benson DA, Bryant SH, Canese K, Chetvernin V, et al. Database resources of the National Center for Biotechnology Information. Nucleic Acids Research. 2007 Jan 1;35(suppl_1):D5–12.

10. Gadin JR, Zetterberg R, Meijsen J, Schork AJ. Cleansumstats: Converting GWAS sumstats to a common format to facilitate downstream applications. 2022.

11. Magnus P, Birke C, Vejrup K, Haugan A, Alsaker E, Daltveit AK, et al. Cohort Profile Update: The Norwegian Mother and Child Cohort Study (MoBa). International Journal of Epidemiology. 2016 Apr 1;45(2):382–8.

12. Refsum H, Nurk E, David Smith A, Ueland PM, Gjesdal CG, Bjelland I, et al. The Hordaland Homocysteine Study: A Community-Based Study of Homocysteine, Its Determinants, and Associations with Disease1. The Journal of Nutrition. 2006 Jun 1;136(6):1731S-1740S.

13. Nygård O, Vollset SE, Refsum H, Stensvold I, Tverdal A, Nordrehaug JE, et al. Total Plasma Homocysteine and Cardiovascular Risk Profile: The Hordaland Homocysteine Study. JAMA. 1995 Nov 15;274(19):1526–33.

14. Willer CJ, Li Y, Abecasis GR. METAL: fast and efficient meta-analysis of genomewide association scans. Bioinformatics. 2010 Sep 1;26(17):2190–1.

15. Mbatchou J, Barnard L, Backman J, Marcketta A, Kosmicki JA, Ziyatdinov A, et al. Computationally efficient whole-genome regression for quantitative and binary traits. Nat Genet. 2021 Jul;53(7):1097–103.

16. Corfield EC, Shadrin AA, Frei O, Rahman Z, Lin A, Athanasiu L, et al. The Norwegian Mother, Father, and Child cohort study (MoBa) genotyping data resource: MoBaPsychGen pipeline v.1. bioRxiv; 2024. p. 2022.06.23.496289.

17. Koenig Z, Yohannes MT, Nkambule LL, Zhao X, Goodrich JK, Kim HA, et al. A harmonized public resource of deeply sequenced diverse human genomes. Genome Res. 2024 Jun 25;34(5):796–809.

18. Smeland OB, Frei O, Shadrin A, O'Connell K, Fan CC, Bahrami S, et al. Discovery of shared genomic loci using the conditional false discovery rate approach. Hum Genet. 2020 Jan;139(1):85–94.
